# ACCIO: An Assembly-Based Tool Enabling Plasmid Detection

**DOI:** 10.1101/2025.10.30.25338662

**Authors:** Nathan J. Raabe, Marissa P. Griffith, Vatsala Rangachar Srinivasa, Kady D. Waggle, Alexander J. Sundermann, Lora Lee Pless, Graham M. Snyder, Maria M. Brooks, Daria Van Tyne, Lee H. Harrison

## Abstract

2.

Plasmids are extrachromosomal mobile genetic elements that often carry genes responsible for antimicrobial resistance. Plasmid epidemiology aims to track the evolution and spread of plasmids, but the field currently faces significant barriers that make practical implementation using whole genome sequence data difficult. Hybrid-assembled genomes remain the most reliable way to identify and track complete plasmids; however, most genomic surveillance data exists in the form of short-read sequencing, which lacks the resolution required to accurately resolve plasmids. Despite recent advances, long-read-only assemblies have not yet reached the consistency seen in hybrid assemblies. The ideal approach to plasmid epidemiology using whole genome sequence data would consider the limitations of sequencing technologies and the constraints of existing genomic surveillance infrastructure, in addition to the unique evolutionary biology of plasmids. Here, we present ACCIO (Assembly-based Circular Contig Identification for Outbreaks), a tool which creates a reference plasmid database and uses it to infer which plasmids, and genetically related plasmid groupings, are present in an input assembly (Illumina, Nanopore, or hybrid assembly). We validated ACCIO using an internal dataset of 303 plasmid-harboring bacterial clinical and surveillance isolates collected from a single acute tertiary care center. When highly related database plasmids were grouped together, ACCIO achieved 100% sensitivity and 92.1% positive predictive value (PPV) for detection of plasmid groups using hybrid assemblies, and comparably strong performance for Illumina (93.0% sensitivity, 86.6% PPV) and Nanopore (79.3% sensitivity, 91.4% PPV) assemblies. Evaluation on three external datasets yielded consistently high performance. Finally, when benchmarked against MOB-suite, a tool for reconstruction and typing of plasmids, ACCIO demonstrated superior performance across nearly all assembly types and plasmid grouping levels. By integrating database construction, clustering, and plasmid calling into a single workflow compatible with all major sequencing platforms, ACCIO is intended to help advance plasmid epidemiology beyond its current technological and infrastructural barriers.

**Impact statement:** Detecting and tracking plasmids—the mobile genetic elements often responsible for spreading antimicrobial resistance in hospital settings—is challenging, particularly when relying on short-read sequencing data alone. Short-read genome assemblies, despite widespread use in surveillance of bacterial pathogens, inherently lack the resolution required for plasmid analyses. Current bioinformatic methods struggle to identify whole plasmids from short-read assemblies alone, and often, hybrid assembly using both short- and long-read data is required for the robust analyses that are essential for tracking plasmids.

To address these challenges, we developed ACCIO, a bioinformatics tool which utilizes input genome assemblies (short-read, long-read, or hybrid assemblies) to assess the plasmid content of clinical bacterial isolates for epidemiologic purposes. We validated its use against the recovery of circular plasmid sequences from hybrid assembled genomes as a gold standard method for determining plasmid content. Using a curated local database of 430 plasmid sequences, ACCIO provided accurate inferences of plasmid content from short-read (Illumina), long-read (Oxford Nanopore Technologies), and hybrid assemblies (both), ultimately facilitating genomic surveillance of plasmids regardless of sequencing technology. This work represents a meaningful step forward in advancing plasmid surveillance beyond the technological and infrastructural barriers that limit its broader expansion into healthcare and other settings.

**Data summary:** Short- and long-read sequencing data have been deposited in the NCBI Sequence Read Archive (SRA) under multiple BioProjects, and corresponding hybrid genome assemblies are available in GenBank. Accession numbers for all BioProjects, BioSamples, and SRA datasets are provided in Supplementary Data S1. All supporting data, software code, and experimental/analysis protocols are provided within the article or in supplementary data files. External validation of ACCIO used three external datasets (Cho et al. 2023, BioProjects PRJNA475751 and PRJNA874473, DOI: 10.1038/s41598-024-70540-1; Lipworth et al. 2024, BioProject: PRJNA604975, DOI: 10.1038/s41467-024-45761-7; Khezri et al. 2021, European Nucleotide Archive (ENA): PRJEB45084, DOI: 10.3390/microorganisms9122560).

*List of External Software:*

1. **MOB-suite (v3.1.9) –** https://github.com/phac-nml/mob-suite
2. **Skani (v0.2.2) –** https://github.com/bluenote-1577/skani
3. **Scipy (v1.16.1) –** https://github.com/scipy/scipy
4. **Pling (v2.0.0) –** https://github.com/iqbal-lab-org/pling
5. **MUMmer / NUCmer (v4.0.1) –** https://mummer4.github.io/
6. **Mash / Mash Screen (v2.3) –** https://github.com/marbl/Mash
7. **SPAdes (v3.15.5) –** https://github.com/ablab/spades
8. **Unicycler (v0.5.1) –** https://github.com/rrwick/Unicycler
9. **Flye (v2.9.5) –** https://github.com/mikolmogorov/Flye
10. **QUAST (v5.2.0) –** https://github.com/ablab/quast
11. **Kraken2 (v2.1.3) –** https://github.com/DerrickWood/kraken2
12. **CheckM (v0.4) –** https://github.com/Ecogenomics/CheckM
13. **Albacore/Guppy –** [no longer officially hosted; was distributed by ONT]
14. **Guppy –** https://nanoporetech.com/software/other/guppy
15. **Dorado –** https://github.com/nanoporetech/dorado
16. **Bowtie2 (v2.5.4) –** https://github.com/BenLangmead/bowtie2
17. **Minimap2 (v2.28) –** https://github.com/lh3/minimap2
18. **Biopython (v1.85) –** https://biopython.org/
19. **Pandas (v2.3.1) –** https://pandas.pydata.org/
20. **Plasme (v1.1) –** https://github.com/HubertTang/PLASMe
21. **BLAST(v2.17.0) –** https://blast.ncbi.nlm.nih.gov/Blast.cgi

## 5. Introduction

Plasmids—circular pieces of extrachromosomal self-replicating DNA—are important drivers of the spread and evolution of antimicrobial resistance (AMR) in healthcare settings (1–3). The bacteria that cause healthcare-associated infections often harbor plasmids which encode genes conferring resistance to commonly prescribed antimicrobial agents—this is often referred to as plasmid-mediated resistance. The result is pathogens that are well adapted to the hospital environment and associated with limited treatment options and poor patient outcomes (4). When implemented, genomic surveillance of plasmids can provide infection prevention teams with opportunities to interrupt the spread of antimicrobial resistance in the hospital setting (5). However, tracking plasmid-mediated resistance for infection prevention and control purposes represents a unique epidemiological challenge due to the mobility of plasmids, in that they can be acquired, carried, and shared between the same or different bacterial species through horizontal transfer (6,7).

Thus, we identify three primary barriers to practical plasmid epidemiology. First, the evolutionary biology of plasmids is different from that of the bacterial chromosome; plasmids exist under distinct selective pressures that shape their persistence, mobility, and genetic compositions (8–12). Plasmids are often, but not always, mobile and plastic; they evolve and move independently of the bacterial chromosome and often lose or acquire genetic material through recombination with, or co-integration of, sequences from the bacterial chromosome or other mobile genetic elements (MGEs) (13–16). The resulting structural changes complicate efforts to define and track plasmid lineages based on measures of genetic similarity that are normally used for genomic epidemiology investigations, such as single nucleotide polymorphism (SNP) comparisons, k-mer sketching (mash distance), or average nucleotide identity (ANI) (17–19).

Second, the horizontal transfer of plasmids among bacterial species in healthcare settings is not accurately detected by current whole genome sequencing (WGS) surveillance approaches, which primarily aim to detect clonal transmission of bacterial species (2,5,20). Horizontal transfer of plasmids can occur within a co-colonized patient or a hospital environment contaminated with more than one bacterial species. These settings where bacterial species are frequently in contact provide opportunities for plasmid transfer between species, which may facilitate transmission of pathogens between hospitalized patients. To better combat the spread of plasmids in hospital settings (21,22), surveillance approaches must operate in real time and accurately infer plasmid content, as focusing only on clonal transmission risks missing plasmid transfer (5,23).

Third, practical applications of plasmid epidemiology are complicated by the inherent limitations of current sequencing technologies. Despite being one of the most widely used types of WGS, short-read sequencing has been associated with challenges in conducting high-resolution plasmid investigations as a result of its inherent limitations (24–28). Due to the presence of repetitive regions in bacterial genomes often exceeding the length of short-reads, short-read sequencing alone struggles to fully resolve structures like plasmids (29– 33). Reconstructing plasmid assemblies from short reads often requires the use of specialized plasmid reconstruction tools, the best of which fail to reliably reconstruct the majority of plasmids carrying ARGs, including large ARG- and ESBL-plasmids (30). Long-read sequencing can generate reads which are long enough to span repetitive regions, resulting in less fragmented genome assemblies more suited for plasmid analyses. However, variable accuracy, persistent error rates even after polishing, and the loss of smaller plasmids remain significant barriers to the widespread adoption of long-read sequencing for plasmid epidemiology (31–34). Hybrid assemblies, which combine short- and long-read data, are highly effective at reconstructing accurate and structurally contiguous genomes, frequently recovering complete, circular plasmids (29,32,35). However, the burden of performing two separate sequencing methods per isolate makes hybrid assembly challenging to implement at the scale required for genomic surveillance networks (36).

In response to these barriers, we developed ACCIO (<u>A</u>ssembly-based <u>C</u>ircular <u>C</u>ontig Identification for <u>O</u>utbreaks), a tool for detecting plasmids in bacterial genome assemblies. ACCIO identifies the presence of highly related plasmids from a curated local plasmid database and determines which plasmid grouping(s) a given assembly most likely contains for downstream investigation of putative plasmid transmission. To account for the complex nature of plasmid evolution, ACCIO leverages tools which quantify the structural features of plasmid evolution (rearrangements, insertions, deletions) in addition to measures of sequence similarity. This approach better defines plasmid groupings to account for the complex nature of plasmid evolution when conducting an epidemiologic investigation. ACCIO allows users to input hybrid assemblies or supply known plasmid sequences to construct a curated reference database that captures both sequence and structural similarity between plasmids—allowing the database to reflect a customized epidemiological context. After establishing a reference database, ACCIO can then be used to evaluate input genome assemblies to infer plasmid content and indicate the presence of plasmid groupings for epidemiologic investigations. To determine which database plasmids and respective plasmid groups are present in an input assembly, ACCIO uses a composite scoring approach that evaluates each assembly’s contigs against the reference plasmid database using multiple metrics and assigns assembly contigs to their best-matching plasmids. ACCIO was developed primarily to address the challenge of plasmid detection in short-read assemblies (Illumina) but is compatible with both long-read (Oxford Nanopore Technologies, ONT) and hybrid assemblies (Illumina + ONT) to offer flexibility across a range of bioinformatic workflows and enable plasmid epidemiology regardless of sequencing technology In this validation analysis, we used ACCIO to construct a local plasmid database containing 430 unique plasmid sequences from the hybrid assemblies of plasmid-harboring clinical isolates collected within a single healthcare system over two decades. We then validated ACCIO’s ability to detect database plasmids and plasmid groupings in the short-read, long-read, and hybrid assemblies of 303 study isolates. The output calls of ACCIO were compared to a ground-truth set of 613 plasmids known to be present via the gold-standard method, based strictly on recovery of circular-and-complete plasmid sequences directly from hybrid assemblies. We found that ACCIO achieved high accuracy in detecting plasmids and plasmid groupings across sequencing platforms, with performance comparable to the hybrid-based ground truth.

## 6. Theory and Implementation

### 6.1 Overview

ACCIO is an open-source, python-based bioinformatics tool developed to support plasmid epidemiology by inferring the plasmid content of bacterial genome assemblies. In practice, ACCIO uses a reference database to determine which plasmids and plasmid groupings are most likely present within a given isolate assembly. The tool operates in two stages: *Stage 1* involves the construction of a reference plasmid database and assignment of groupings to database plasmids based on genetic similarity, and *Stage 2* entails the comparison of input genome assemblies against the database to determine which plasmids and plasmid groupings are present or absent within an assembly.

### 6.2 ACCIO Stage 1A: Creation of a curated local plasmid database from hybrid assemblies and/or known plasmid sequences

#### Supplying a reference plasmid database

ACCIO requires a plasmid reference database from which plasmids and plasmid groupings can be chosen. In *Stage 1*, ACCIO creates and processes the plasmid database used later in *Stage 2* for inferring plasmid content in assemblies (**Fig. 1A**). The database can be constructed from known plasmid sequences or can be populated by ACCIO from a set of input hybrid-assembled genomes. If hybrid assemblies are used as input, plasmids are identified by ACCIO as circular and complete contigs >10kbp in length which encode plasmid replicons identifiable by BLASTn (≥80% similarity score) to the PlasmidFinder database or using MOB-typer(37,38). Circularity information is obtained directly from hybrid assembly contig headers, or if contig ends overlap by at least 25 bp.

**Fig. 1.**
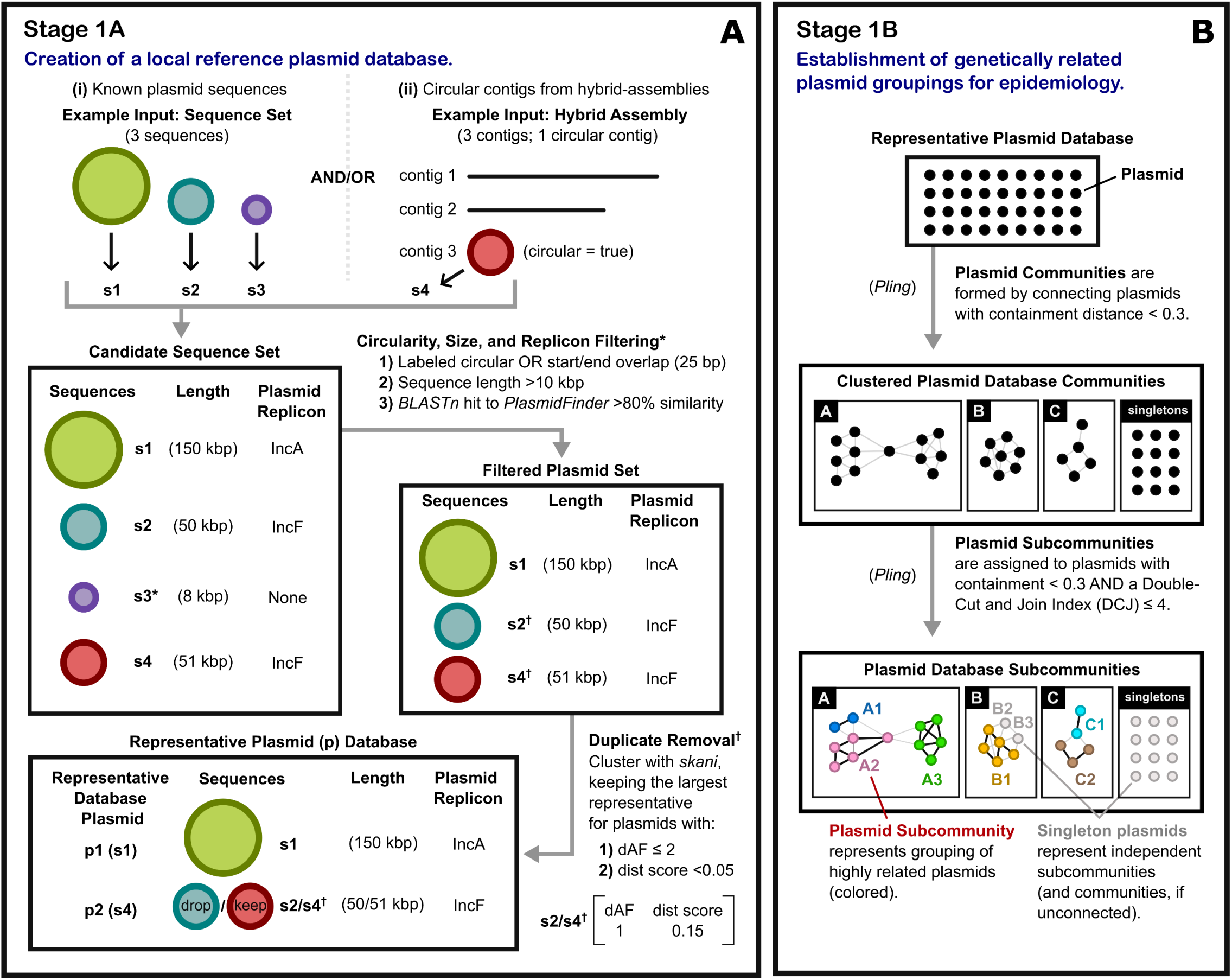
ACCIO Stage 1: Construction of a reference plasmid database and assignment of plasmid groupings. **A)** In Stage 1A, ACCIO creates a curated local reference plasmid database using an inputted set of hybrid assemblies or known plasmid sequences. **B)** In Stage 1B, ACCIO uses Pling to assign database plasmids into plasmid communities and subcommunities based on genetic similarity. Kbp, kilobase pair; dAF, alignment fraction distance; dist score, combined distance score. Examples: * s3 removed based on length (<10 kbp) and lack of plasmid replicon. † s2 and s4 met similarity thresholds (dAF, dist score); longer sequence (s4) retained as the representative plasmid (p2).

#### Removing database redundancy

A crucial step in the creation of the plasmid reference database is clustering plasmid sequences based on genetic similarity to remove redundancy and keep only representative plasmids. ACCIO utilizes skani to calculate pairwise alignment fraction (AF) and ANI between each plasmid in the filtered database, which are then used to calculate distance values: average nucleotide identity distance (dANI = 100 – %ANI) and minimum alignment fraction distance (dAF = 100 – min(%AF₁, %AF₂)) (39). A combined distance score is then calculated as ([0.5 × dANI] + [0.5 × dAF / 40]). Pairs of plasmids with <2 minimum dAF (or >98% AF) are then filtered out to avoid instances of pairs with high nucleotide identity but poor alignment fraction. A distance matrix is created, and average linkage clustering is performed using the combined distance score with a threshold of <0.05 (scipy)(40). A representative plasmid is chosen from each resulting cluster as the longest plasmid in the cluster, and all other plasmids within the cluster are considered duplicates and removed from the database. The final deduplicated reference database consists of only unique plasmids and representative plasmid sequences from each cluster of genetically related sequences.

### 6.3 ACCIO Stage 1B: Establishment of genetically related plasmid groupings for epidemiology

#### Plasmids, plasmid subcommunities, and plasmid communities

To enable meaningful downstream epidemiologic analyses, such as tracking plasmid dissemination, identifying transmission clusters, and assessing the spread of resistance genes, each plasmid in the representative reference database is assigned to both a *plasmid community* (a coarse grouping indicating broad genetic relatedness based on lower-resolution similarity metrics) and a *plasmid subcommunity* (a refined grouping reflecting close relatedness and potential plasmid transmission clusters) using Pling (**Fig. 1B**) (41). Pling first clusters plasmids into *plasmid communities* by building a network between them based on the amount of shared sequence; this is accomplished using a containment distance threshold (the proportion of the smaller plasmid that is not contained in the larger plasmid) which is calculated from pairwise alignments with NUCmer (42). Aligned regions are then transformed into ordered blocks using integerization, where each integer represents a gene or syntenic block with orientation (positive or negative). The integerized plasmids are then compared using the Double Cut and Join with Indels (DCJ-Indel) Model to estimate the number of structural variations between two plasmids—rearrangements, insertions, and deletions—which would be required to transform one plasmid into another. Because plasmids evolve through frequent structural changes, the resulting DCJ-Indel distance provides a more informative measure of evolutionary divergence between plasmids than sequence similarity-based methods alone. For epidemiological applications, it is recommended that a conservative containment distance threshold of <0.3 and DCJ-Indel distance of ≤4 are used to connect plasmids which are similar enough to represent possible recent transmission events—meaning that for two plasmids to be part of the *same community*, at least 70% of the smaller plasmid must be contained in the larger plasmid, and for two plasmids to be part of the *same subcommunity*, they must also have a DCJ-indel distance of ≤4 structural variations between them (41). After edges between plasmids are established based on containment and DCJ thresholds, Pling determines subcommunity assignments based on an asynchronous label propagation network community algorithm which also considers connections to hub plasmids (plasmids identified by Pling as densely connected plasmids whose neighbors are sparsely connected to one another). Because hub plasmids often contain transposable elements or MGEs common to multiple plasmids and connect otherwise unrelated groups, ACCIO treats them as independent subcommunities.

#### Flexibility for connections between large plasmids

Based on the results of a multivariable-adjusted linear regression model describing the relationship between DCJ-Indel distance and plasmid size among our database plasmids, we observed that DCJ-Indel distance increases with increasing plasmid size adjusting for containment distance (**Fig. S1**). To account for this, ACCIO creates plasmid subcommunities using a DCJ threshold of ≤4 whenever the smaller plasmid in a pairwise comparison is ≤120 kbp and allows +1 DCJ for every 20 kbp range above this threshold. Ultimately, these parameters can be changed by the user to allow for more exclusive or inclusive groupings. For the present study, we applied more stringent cutoffs to better reflect the requirements of epidemiological analyses, where high specificity and positive predictive value are critical for accurately identifying plasmid transfer events.

### 6.4 ACCIO Stage 2A: Identifying plasmid replicons and repetitive contigs in assemblies

#### Assembled contigs as a basis for plasmid inferences

The primary input for *Stage 2* of ACCIO is a genome assembly in the form of a fasta file, which serves as a basis for inferring which plasmid(s) and plasmid grouping(s) are present from the plasmid database constructed in *Stage 1*. In the case of assemblies which are fragmented into many small contigs, like those produced using short-read sequencing data, it is unlikely that a plasmid will be represented as a single circular contig, as often occurs in hybrid assemblies, or somewhat less frequently, in long-read-only assemblies. ACCIO matches contigs from short-read, long-read, or hybrid assemblies to database plasmids and ultimately calls plasmids using a composite scoring system that incorporates multiple facets of genetic relatedness. If the raw sequencing reads are provided in addition to the genome assembly (recommended for short-read-only assemblies), ACCIO also calculates additional metrics using raw reads and applies these metrics towards the composite score.

#### Plasmid replicon type filtering

Before scoring contigs, ACCIO first performs BLASTn to compare the input isolate assembly against the PlasmidFinder database to identify the plasmid replicon families (i.e., IncFII, not IncFII(X)) present in the assembly (>80% similarity score ([(% identity) × (% coverage)] / 100)). The reference plasmid database is then filtered to include only plasmids with replicon types which fall under the broader replicon families identified in the base assembly (**Fig. 2A**). From this point forward, contigs in the input assembly are then scored against the filtered set of database plasmids which have replicon families also known to be present in the base assembly.

**Fig. 2.**
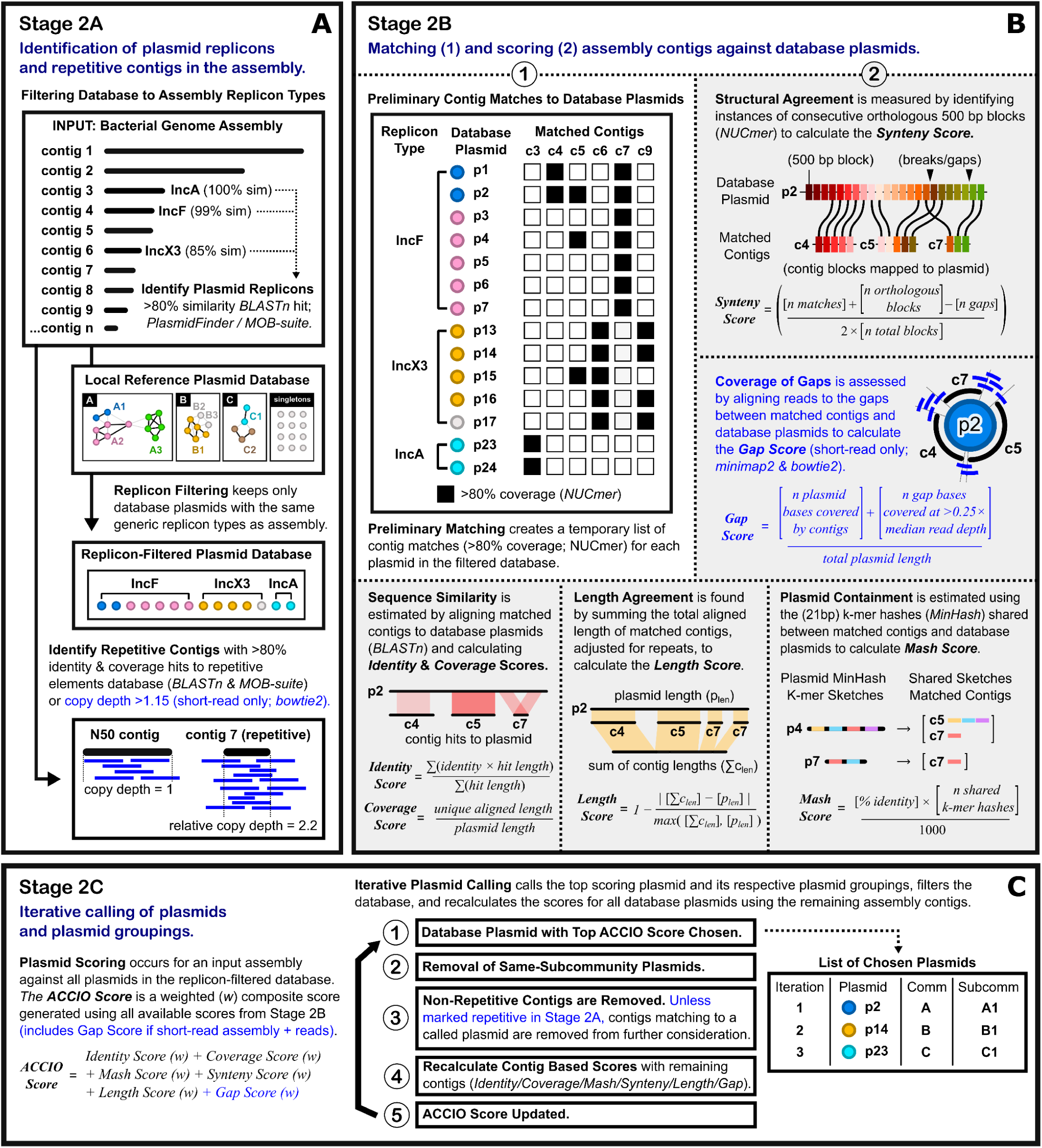
ACCIO Stage 2: Comparison of input genome assemblies against the database to call plasmids and plasmid groupings. **A)** In Stage 2A, ACCIO identifies plasmid replicons and repetitive contigs in the input genome assembly. **B)** In Stage 2B, ACCIO matches and scores assembly contigs against database plasmids. Contigs are first matched to database plasmids using NUCmer—the set of matching contigs is then used to generate the Identity, Coverage, Mash, Synteny, Length, and Gap Scores. **C)** In Stage 2C, ACCIO iteratively calls plasmids and their corresponding communities and subcommunities using the composite ACCIO Score. Blue text indicates processes specific to short-read input by default. p, plasmid; c, contig; subcomm, subcommunity; comm, community.

#### Identification of contigs representing repetitive elements

Due to the repetitive regions found among plasmid sequences, assembly contigs which represent or contain these regions could plausibly belong to multiple plasmids within the database, particularly when bacteria harbor more than one plasmid. To identify repetitive assembly contigs, BLASTn is performed against the ‘repetitive elements database’ from MOB-suite (38). If a contig contains a hit (>80% BLASTn identity and coverage) to an element in the database, it is flagged as repetitive. If sequencing reads are provided, the relative copy depth for each assembly contig is calculated by aligning reads back to the assembly (bowtie2) to estimate the median read depth across each contig (43). ACCIO uses a method for calculating copy depth described by Unicycler; briefly, the mean depth of the N50 contig (the median contig selected from the 10 longest assembly contigs) is used as a reference, and copy depth for each contig is calculated ([mean contig depth] / [mean depth of N50 contig]) (44). Contigs with >1.15 copy depth are flagged as repetitive. When final assignment of contigs to database plasmids occurs based on the scores generated by ACCIO in later steps, repetitive contigs are allowed to match to more than one chosen plasmid, while non-repetitive contigs may only match to one database plasmid.

### 6.5 ACCIO Stage 2B: Matching and scoring assembly contigs against database plasmids

#### Preliminary matching of contigs to database plasmids

Assembly contigs are first compared to each plasmid in the replicon type-filtered reference database using NUCmer, a pairwise alignment tool (**Fig. 2B**) (45). Contig alignments with >80% coverage to a database plasmid are then preliminarily ‘matched’ to that plasmid—this step is conducted for each plasmid in the database, and a set of the contig matches for each assembly-plasmid comparison is created and used downstream for all other score calculations.

#### Measuring structural agreement between matched contigs and plasmids

Because of the large structural component of plasmid evolution, such as rearrangements of genes and other genomic regions, inversions, and gene gain or loss, ACCIO quantifies the synteny, or structural similarity, between assembly contigs and database plasmids using NUCmer, which can be used to capture structural divergence between sequences by identifying matching sequence “blocks” (45). In practice, NUCmer results are filtered to retain the best hits between each database plasmid and its set of matched contigs (no small overlapping hits, one hit per plasmid region). The database plasmid and its set of matched assembly contigs are then divided into 500 base pair blocks. Each block is considered orthologous if the next consecutive block maps to the same contig in the input assembly or to a new contig not yet mapped in the results. If the next consecutive block instead maps back to a different contig already in the results, this is considered a break/gap. From this, a *synteny score* is calculated as ([(number of matches) / (number of total blocks)] + [(number of orthologous blocks) / (number of total blocks)] – [(number of gaps) / (number of total blocks)]). The *synteny score* quantifies the level of structural conservation between the input assembly contigs and database plasmids.

#### Coverage of gaps between contigs matched to plasmids in short-read assemblies

Genome assemblies made using short reads are often fragmented into dozens to hundreds of contigs. To address this limitation, when determining the plasmid content of an input short-read assembly, ACCIO uses raw reads (if provided) to assess the coverage of gaps between the assembly contigs that are matched to the same database plasmid. Gap regions of the plasmid are identified from the NUCmer hits of the matching contigs (42). Short reads are then aligned to a database plasmid (bowtie2) (43), which assigns each read to its best-scoring alignment location, and alignment depths are calculated for each position of the plasmid sequence. An additional scoring metric for short read assemblies, the *gap score*, is then calculated for each plasmid-contig set by obtaining the number of gap bases covered (>0.25 × the median read depth).

#### Quantifying sequence similarity between matched contigs and plasmids

To quantify the sequence similarity between assembly contigs and database plasmids, BLASTn is used to align the set of matched contigs against their corresponding database plasmids to calculate *BLASTn identity and coverage scores*. Results are filtered to only the best hits for each contig, as defined by: 1) ≥75% identity to a database plasmid, 2) ≥75% coverage of the contig OR a hit spanning the beginning or end of the contig, and 3) not enveloped (shorter, lower-quality hits contained entirely within a larger, higher-scoring hit on the same contig). Using the set of filtered hits from matched contigs, a *BLASTn identity score* (weighted identity from all filtered BLASTn hits from matched contigs to plasmid) and *BLASTn coverage score* (% coverage from all filtered BLAST hits from matched contigs to plasmid) are calculated, which together represent sequence-level similarity between the matched contigs and each database plasmid.

#### Determining agreement between lengths of plasmid and matched contigs

To determine how closely the length of matching contigs approximates that of the reference plasmid, we define a *length score* (1 – ([|∑ contig lengths − plasmid length|] / [max(∑ contig lengths, plasmid length)])) based on the total aligned length of all contigs matched to a given plasmid. Non-repetitive contigs are represented once while contigs marked as repetitive in *Stage 2A* are counted towards the length score according to their copy depth relative to the average depth of non-repetitive contigs matched to the same plasmid ([repetitive contig length] × [min([repetitive contig depth] / [avg non-repetitive contig depth], [number of times contig is covered at >80% in the plasmid based on NUCmer alignments]. Thus, the *length score* represents the agreement between the length of the matched contigs from the assembly, considering repetitive contigs, and the length of the corresponding reference plasmid.

#### Estimating plasmid containment using matched contigs

Mash Screen, an alignment-free algorithm which measures the containment of genomes within assembled sequences or sequencing reads, is run for each set of matched contigs against the corresponding database plasmid. A *mash containment score* is computed as ([(% identity) × (number of shared MinHash k-mer hashes)] / 1000) (46). Shared hashes represent overlapping k-mers (default 21 bp) found in both the reference plasmid and the query input, and thus, the *mash score* itself represents how much of the reference plasmid is contained within the query. Mash is the metric used in MOB-suite for identifying a nearest database plasmid (mash_nearest_neighbor) to a given plasmid sequence (38,46,47). Similarly, ACCIO also uses mash containment as one metric to help assess which plasmid(s) from a database are most likely to be present in an assembly.

### 6.6 ACCIO Stage 2C: Iterative calling of plasmids and plasmid groupings

#### Calling plasmids and groupings using ACCIO’s composite score

ACCIO calculates a composite score (*ACCIO Score*) for each plasmid in the filtered database as a weighted sum of the scores generated in *Stage 2B* (**Fig. 2C**). Weights were empirically chosen based on repeat testing using our validation dataset to balance the contribution of each component score and optimize overall classification performance. When the input is a short-read assembly and raw short reads are provided, the weighted *ACCIO Score* is calculated as *BLASTn identity score* (×0.09) + *BLASTn coverage score* (×0.16) + *mash score* (×0.24) + *synteny score* (×0.12) + *length score* (×0.17) + *gap score* (×0.22). For all other inputs (short-read assemblies without raw reads, long-read assemblies, and hybrid assemblies) the *ACCIO Score* is calculated as *BLASTn identity score* (×0.13) + *BLASTn coverage score* (×0.20) + *mash score* (×0.26) + *synteny score* (×0.17) + *length score* (×0.24) + *gap score* (×0). The database plasmid with the highest *ACCIO Score* is called as present in the genome assembly, along with its corresponding subcommunity and community designated in *Stage 1B*. For the plasmid with the highest *ACCIO Score* to be called as present, the set of matched assembly contigs must have a minimum *mash score* greater than the default threshold (*mash score* ≥ 0.996), which was selected to optimize the removal of false positives with minimal loss of true positives (**Fig. S2**). This *mash score* threshold can be adjusted by the user (suggested range: 0.994-0.998) to prioritize sensitivity (lower values) or PPV (higher values).

#### Iterative plasmid calling with stepwise pruning of non-repetitive contigs and redundant database plasmids

Once the database plasmid with the highest *ACCIO Score* is called as present, the process is repeated iteratively with additional filtering steps to continue calling plasmids based on the remaining assembly contigs: 1) other database plasmids from the same subcommunities as previously called plasmids are removed from further consideration, 2) unless marked as repetitive in *Stage 2A*, contigs matching to previously called plasmids are excluded for all subsequent iterations, 3) *identity*, *coverage*, *mash*, *synteny*, *length*, and *gap scores* are then recalculated for each database plasmid using the remaining contigs, 4) the overall score is then updated for all remaining plasmids before the next highest scoring plasmid is called. The process of calling the highest-scoring plasmid, removing plasmids from the same subcommunity, excluding contigs matched to called plasmids from subsequent comparisons, and recalculating scores is repeated iteratively until no assembly contig set exceeds the mash score threshold for its matched database plasmid or until no contigs or database plasmids remain.

#### Identification of novel plasmids

After calling database plasmids, ACCIO uses remaining contigs to identify instances where an isolate assembly likely contains a novel plasmid not yet represented in the database. Contigs are first classified as plasmid or non-plasmid; contigs are designated as plasmid if they contain an identifiable plasmid replicon (>80% similarity PlasmidFinder hit) or are assigned as plasmidic by PLASme, a machine learning–based classifier that distinguishes plasmidic from chromosomal sequences using sequence composition and homology (48). The resulting plasmid contigs are then provided to mob_recon as a single input file, and plasmids are reconstructed using MOB-suite’s default database as reference (38). Plasmids reconstructed by mob_recon and the assembly contigs used to generate them are reported as novel plasmids alongside ACCIO’s calls from the local database.

### 6.7 Study isolates and setting

Study isolates comprising the complete dataset used to construct a plasmid database and evaluate the performance of ACCIO (N = 532 isolates) were collected from patient cultures obtained for either clinical care or surveillance purposes, primarily at UPMC Presbyterian Hospital, an adult tertiary care facility with 699 beds, 134 critical care beds, and over 400 solid organ transplantations annually. The majority of these isolates (n = 402, 75.6%) were collected from November 2016 to May 2024 as part of EDS-HAT (the Enhanced Detection System for Healthcare-Associated Transmission), a whole genome sequencing surveillance system which collects select high-priority healthcare-associated pathogens from patients with i) hospital admission dates greater than 2 days before the culture date, or ii) inpatient or outpatient encounters at any UPMC facility in the 30 days prior to the culture date (49). All other isolates were obtained either through UPMC Infection Prevention and Control investigations and sequenced by the Microbial Genomics Epidemiology Laboratory (MiGEL) (n=107, 20.1%; culture dates 2008–2024) or were isolates with antibiotic resistance profiles of interest (n = 23, 4.3%; culture dates 2004– 2023).

### 6.8 Microbiologic methods

Isolates were subcultured onto blood agar plates and grown overnight at 37°C in preparation for DNA extraction and whole genome sequencing.

### 6.9 Whole genome sequencing and assembly

DNA extraction was performed using a single colony taken from overnight cultures and the MagMax DNA Multi-Sample Ultra 2.0 kit on a KingFisher Apex instrument (Thermo Fisher Scientific) or DNeasy Blood and Tissue Kit (Qiagen). Whole genome sequencing for study isolates was performed using both Illumina and Oxford Nanopore Technologies (ONT) platforms.

*Short-read sequencing* was performed on the Illumina MiSeq, NextSeq 550 or 1000, or NovaSeq X Plus platform using an Illumina DNA Prep Tagmentation Kit on an EpMotion and libraries were sequenced using 2 × 150 bp paired-end reads (v2.5 300-cycle kit). Samples were demultiplexed using bcl2fastq v2.20 and Illumina short-read-only assemblies were created using either SPAdes v3.15.5 (50) or Unicycler v0.5 (44).

*Long-read sequencing* was performed using an Oxford Nanopore Technologies MinION Mk1C device with either R9.4.1 or R10.4.1 flow cells and the SQK-RBK004 or SQK-RBK114.24 rapid gDNA barcoding kits. Basecalling and demultiplexing were performed for Nanopore sequencing data using Albacore v2.3.3 (default parameters), Guppy v2.3.1/v6.3.9 (default parameters), or Dorado v0.9.6 (high accuracy model; dna_r10.4.1_e8.2_400bps_hac@v5.0.0). Nanopore-only assemblies were created using Flye v2.9.4 (51). Hybrid assembly using both Illumina and Nanopore sequencing data was performed for all isolates with both short- and long-read data available using Unicycler v0.5.0 (44).

*WGS data quality control* was conducted using QUAST v5.2.0 (52), and samples with less than 35× and 10× average coverage depth for Illumina and Nanopore, respectively, were not included in the test dataset. Species-level classification of reads was performed using Kraken2 v2.12.2 (53) with the standard Kraken database. Isolates passed quality control if the most abundant species identified by Kraken matched the expected species, the assembly length was within 20% of the expected genome size for the species, and the total number of contigs was ≤350. In addition, CheckM (54) was run for all species to check for contamination, and isolate assemblies showing over 5.0% contamination were removed.

### 6.10 Validation analysis methods

#### Datasets and ground-truth definition

Validation analyses to measure the performance of ACCIO were conducted using multiple datasets consisting of isolates with all three assembly types available (Illumina, Nanopore, and hybrid). The *internal validation dataset* was the primary set used to assess the performance of ACCIO and consisted of study isolates where ≥1 plasmid could be identified in the hybrid assembly as a circular-and-complete plasmid contig with an identifiable plasmid replicon. The same approach was applied to *three external datasets*, each consisting of isolates which met the same criteria. This method of recovering plasmids from hybrid assemblies was considered the gold-standard for this analysis. For each isolate in a given dataset, an isolate’s *ground-truth set of plasmids* refers to the plasmids known to be present in that isolate’s hybrid assembly. The total *ground-truth count in a dataset* represents the aggregate number of plasmids recovered from the hybrid assemblies of all isolates in that dataset.

#### Classification counts

To evaluate the performance of ACCIO in correctly calling (classifying) plasmids as present given a study isolate’s input assembly, we defined performance metrics based on comparisons between the set of plasmid calls made by ACCIO and the ground-truth set of plasmids. This analysis was performed at the plasmid, subcommunity, and community levels (using the grouping assignments of ground-truth plasmids and plasmids called by ACCIO). For example, at the plasmid level, positive and negative calls refer to the presence or absence of individual plasmids per isolate as called by ACCIO. For a given isolate, true positives (TP) represent correct positive calls, where a plasmid was present in both the set of plasmids called by ACCIO and the ground-truth set of plasmids for that isolate. False negatives (FN) represent missed calls, where a plasmid was present in the ground-truth set but not the set called by ACCIO. False positives (FP) represent additional calls, where a plasmid called by ACCIO was not present in the ground-truth set of plasmids. True negatives (TN) represent correct negative calls, defined as the number of database plasmids which were not present in both the ground-truth set and the set of plasmids called by ACCIO for a given isolate, calculated as [number of database plasmids] – [TP+FP+FN] for each isolate.

#### Performance characteristics

Overall performance characteristics were calculated for the entire dataset using the aggregate TP, FN, FP, and TN counts for each assembly input type. For example, at the plasmid level, sensitivity (also known as recall) was defined as the proportion of truly present plasmids that were correctly called as present (TP/[TP+FN]); specificity was defined as the proportion of truly absent plasmids that were correctly not called (TN/[TN+FP]); positive predictive value (PPV, also known as precision) was defined as the proportion of plasmids called as present that were truly present (TP/[TP+FP]); negative predictive value (NPV) was defined as the proportion of plasmids not called that were truly absent (TN/[TN+FN]); Jaccard Index was defined as the proportion of overlap between plasmids ACCIO calls as present and plasmids truly present (TP/[TP+FP+FN]); and F1 Score was defined as the harmonic mean of recall and precision (2×TP/[2×TP + FP + FN]). The F1 score provides a single summary metric for test performance, capturing the balance between sensitivity and PPV. Significance testing for differences in sensitivity and PPV were conducted using two-sample tests for equality of proportions with Chi-square approximations

#### Methods for comparison with a current standard toolkit

To assess ACCIO’s performance relative to that of an established tool, we utilized MOB-suite to detect database plasmids in isolate assemblies from the internal validation dataset and compared the results. First, MOB-cluster was run on the plasmid database generated by ACCIO using default parameters. MOB-recon was then run for all isolate assemblies in the validation dataset to obtain a set of reconstructed plasmids which were putatively present in each assembly type. Circular contigs were marked and processed accordingly by enabling the option to check for a circularity flag generated by Unicycler in fasta headers. The nearest mash neighbor plasmid identified by MOB-typer, which represents the closest database plasmid to each MOB-recon plasmid by mash distance, was considered to be MOB-suite’s chosen plasmid for this analysis. Using the community and subcommunity information (obtained via Pling) for each plasmid chosen by MOB-suite, performance was calculated at each group level. Because Pling accounts for structural variation (DCJ-Indel distance metric) in addition to containment, Pling clustering was chosen over MOB-cluster’s primary and secondary cluster assignments to obtain more comparable results.

#### Datasets and methods for supplemental analyses

We also evaluated the performance of ACCIO given Illumina- and Nanopore-only assembly inputs and compared to the results to that of ACCIO using hybrid assemblies. This analysis used the full initial set of study isolates with all three assembly types available, including those without any circularized plasmids in the hybrid assembly. For this analysis, the set of plasmids called by ACCIO using the hybrid assemblies of each isolate served as the set of ground-truth plasmids. As part of another supplemental analysis which used the internal validation set, we assessed whether the plasmid replicons identified in either the set of ground-truth plasmids recovered from hybrid assemblies or those in the set of plasmids called by ACCIO better represented the plasmid replicons identified in base isolate assemblies (standard of comparison). Identifiable plasmid replicons were hits with ≥80% similarity ([% identity * % coverage] / 100) in the PlasmidFinder database.

## 7. Results / Discussion

### 7.1 A local reference plasmid database created from hospital isolates

#### Plasmid database creation and filtering

We constructed a curated local reference plasmid database by screening the hybrid assemblies of 500 clinical and surveillance isolates collected over two decades for unique plasmids (**Supplementary Data S1;** Isolate Metadata). During the database creation step (ACCIO Stage1A), 1033 circular contigs were recovered from the hybrid assemblies of 342 of the 500 isolates, and this set of sequences was supplemented with 53 previously collected, known plasmid sequences from 32 additional isolates for which hybrid assemblies were unavailable. At this step, the initial database contained 1086 circular sequences from a total of 374 isolates before any additional filtering. To retain only sequences which represented plasmids, 423 of the 1086 sequences which had a length <10kbp (98.8%, 418/423) and/or lacked identifiable plasmid replicons (1.2%, 5/423) were filtered from the database by ACCIO, leaving 663 plasmid sequences in the initial database. Very small, highly related plasmids often contain a limited amount of unique sequence, making discrimination between them difficult. Consequently, ACCIO’s detection range is limited to plasmids ≥10 kbp by default. However, among the 339 of 418 plasmids that had an identifiable replicon and lengths below the size cutoff, fewer than 5% (15/339) harbored an ARG (≥80%-similarity hit in the AMRfinder+ database), suggesting that few clinically and epidemiologically relevant AMR-encoding plasmids were lost to database filtering steps. Using ACCIO’s default thresholds for identification of duplicates, 233 of the 663 plasmids were removed from the filtered database, leaving a final plasmid database which consisted of 430 unique, representative plasmid sequences from a diverse set of replicon families (including IncB/C/F/H/I/K/L/M/N/P/R/U/X/Y/Z, Col, and other replicon groups) (**Supplementary Data S1;** Plasmid Metadata). The majority of database plasmids (285/430, 66.3%) carried at least one ARG (≥80% similarity hit to AMRfinder+ database). Among plasmids carrying at least one ARG, the median number of ARGs per plasmid was six and the range was 1-23.

#### Rationale and limitations regarding a local plasmid database

Compared to large public databases containing tens of thousands of sequences, the curated local reference database we constructed with ACCIO provided a collection of plasmids inherently more representative of the plasmids present in our hospital population. While more comprehensive databases, such as MOB-suite’s database (which includes over 20,000 plasmids), offer broader species and global geographic coverage, the scales of such databases may introduce matches to more distantly related plasmids or plasmids with wide geographic distributions, which might not be specific enough to be useful for local epidemiology (38). When making inferences regarding the plasmid content of genome assemblies from clinical isolates, the use of a local plasmid database improves the interpretability and relevance of ACCIO’s output by searching for plasmids already known to exist and circulate locally. One practical limitation of ACCIO is that many settings using a single sequencing technology may not possess the resources required to generate a plasmid database from local hybrid assemblies. In such cases, we recommend either 1) building a database from publicly available plasmid sequences of concern, prioritizing sequences suspected to be circulating locally, or 2) performing targeted sequencing of isolates with novel plasmids and concerning AMR profiles which lack matches in the initial database. The latter strategy may represent a feasible long-term solution for surveillance systems primarily using a single sequencing technology, where the local database may be periodically updated as new plasmids are identified and recovered. As long-read technology and assembly algorithms improve, routine recovery of circular plasmids from long-read-only data may allow for local databases to be populated and updated without the need for hybrid assembly.

#### Plasmid database community and subcommunity compositions

The local plasmid database we constructed clustered the 430 database plasmids into 79 communities, including 49 singleton communities (62.0%) and 30 communities with ≥2 plasmids (38.0%, median size n=4, range 2-222 plasmids) (**Fig. 3, Fig. S3**). Within these communities, the database was further divided into 246 subcommunities, including 189 singleton subcommunities containing only one plasmid (76.8%) and 57 subcommunities with ≥2 plasmids (23.2%, median size n=3, range 2-18 plasmids). Because community and subcommunity designations rely on measures of genetic similarity (containment and DCJ Index) rather than replicon typing, plasmid replicon types are not always homogenous within a given subcommunity. Subcommunities sometimes contained multiple plasmid replicon types due to instances of highly-similar sequences shared across both single replicon and mosaic plasmids, likely resulting from recombination or co-integration events between plasmids of different replicon types over time (15,55). The largest community, Community A, contained 94 subcommunities made up of 222 plasmids recovered from seven gram-negative Enterobacterales species groups (including *Citrobacter* spp.*, Enterobacter hormaechei, Escherichia coli, Klebsiella* spp.*, Proteus mirabilis, Raoultella planticola,* and *Serratia marcescens*). These plasmids formed a sprawling network of genetic relatedness with extensive instances of shared sequences amongst plasmids and across subcommunities. Enterobacterales are recognized as promiscuous hosts of plasmids which are frequently involved in conjugative transfer of plasmids, likely explaining the vast interconnectedness observed within this community (56). Other communities either consisted of plasmids recovered from a single species (Communities B and C were both *Enterococcus faecium* plasmids; Communities D contained *K. pneumoniae* plasmids; Communities E, F, and H contained *E. coli* plasmids) or from multiple Enterobacterales species.

**Fig. 3.**
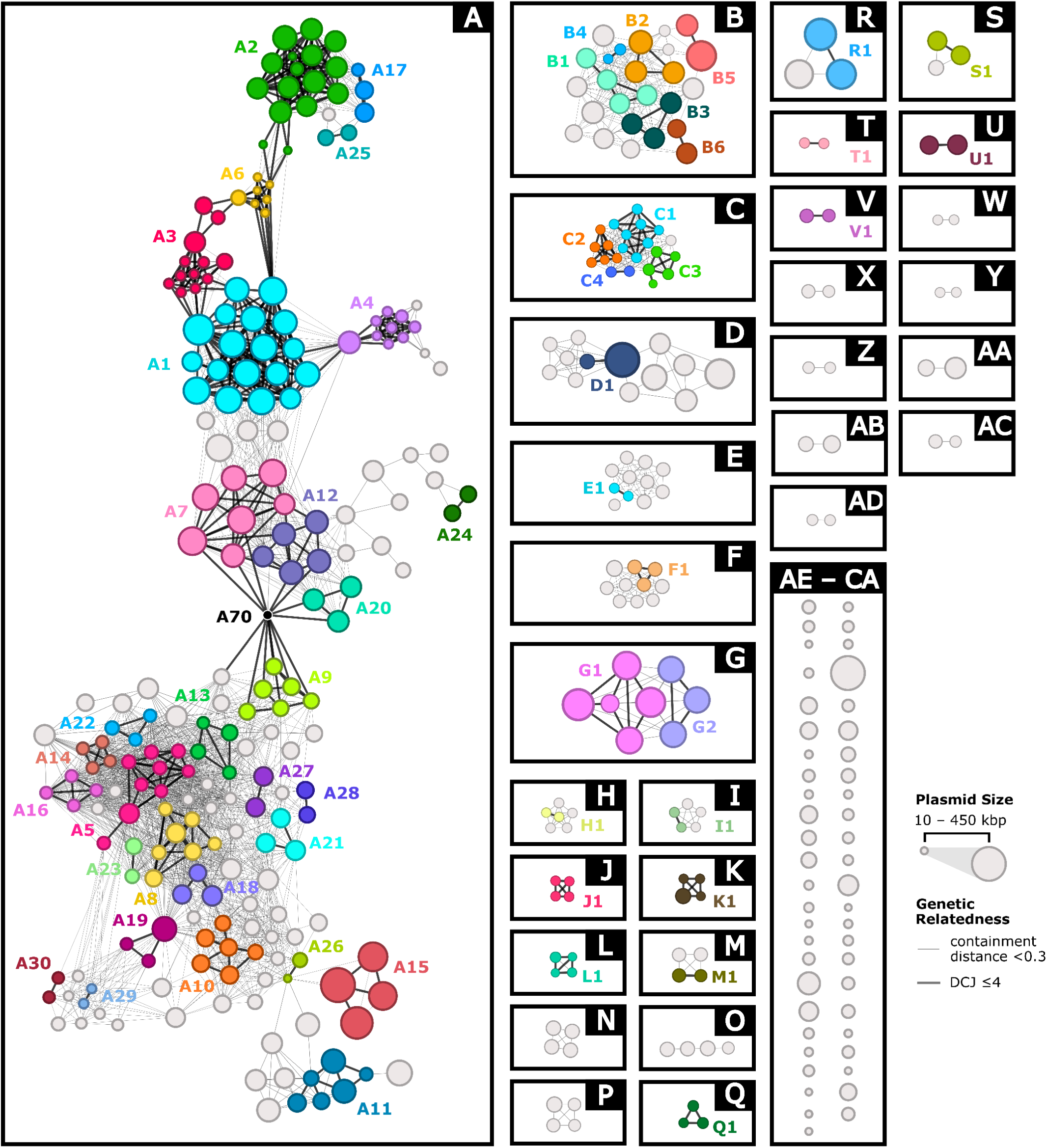
Clustered local reference plasmid database created using ACCIO. The local reference plasmid database (n=430 representative plasmids) was clustered using Pling to define plasmid communities (boxed, labeled A–CA by descending size) based on a containment distance threshold of <0.3. Within each community, subcommunities (colored groups, labeled as community letter plus subcommunity number for all subcommunities containing ≥2 plasmids) represent groups of closely related plasmids defined by high structural similarity (DCJ-Indel distance ≤4 for plasmids <120 kbp, +1 DCJ per additional 20 kbp). Light grey edges represent links between plasmids below the containment distance threshold; black edges represent links between plasmids whose containment distance and DCJ distance are both below the thresholds for relatedness. Plasmid nodes are scaled by size (∼10–450 kbp). Light grey nodes represent plasmids which lacked strong relatedness to any other plasmid in the database and thus were assigned to their own subcommunity. A black node denotes a hub plasmid found in Community A, treated here as its own subcommunity.

### 7.2 Performance assessment using the internal validation dataset

#### Validation dataset and ground-truth counts

The *internal validation dataset* consisted of 303 study isolates for which ≥1 plasmid could be identified in the hybrid assembly as a circular-and-complete plasmid contig with an identifiable plasmid replicon. For each isolate in the *validation dataset*, we compared the set of database plasmids, subcommunities, and communities called by ACCIO to those identified in the *ground-truth set of plasmids* for that isolate and calculated performance characteristics across all three assembly input types for the entire dataset (**Fig. 4**, **Table 1**. Dataset). In total, 613 database plasmids, 604 subcommunities, and 466 communities were identified in the hybrid assemblies of validation set isolates (*ground-truth counts*). Performance varied by assembly input type and by plasmid grouping levels.

**Fig. 4.**
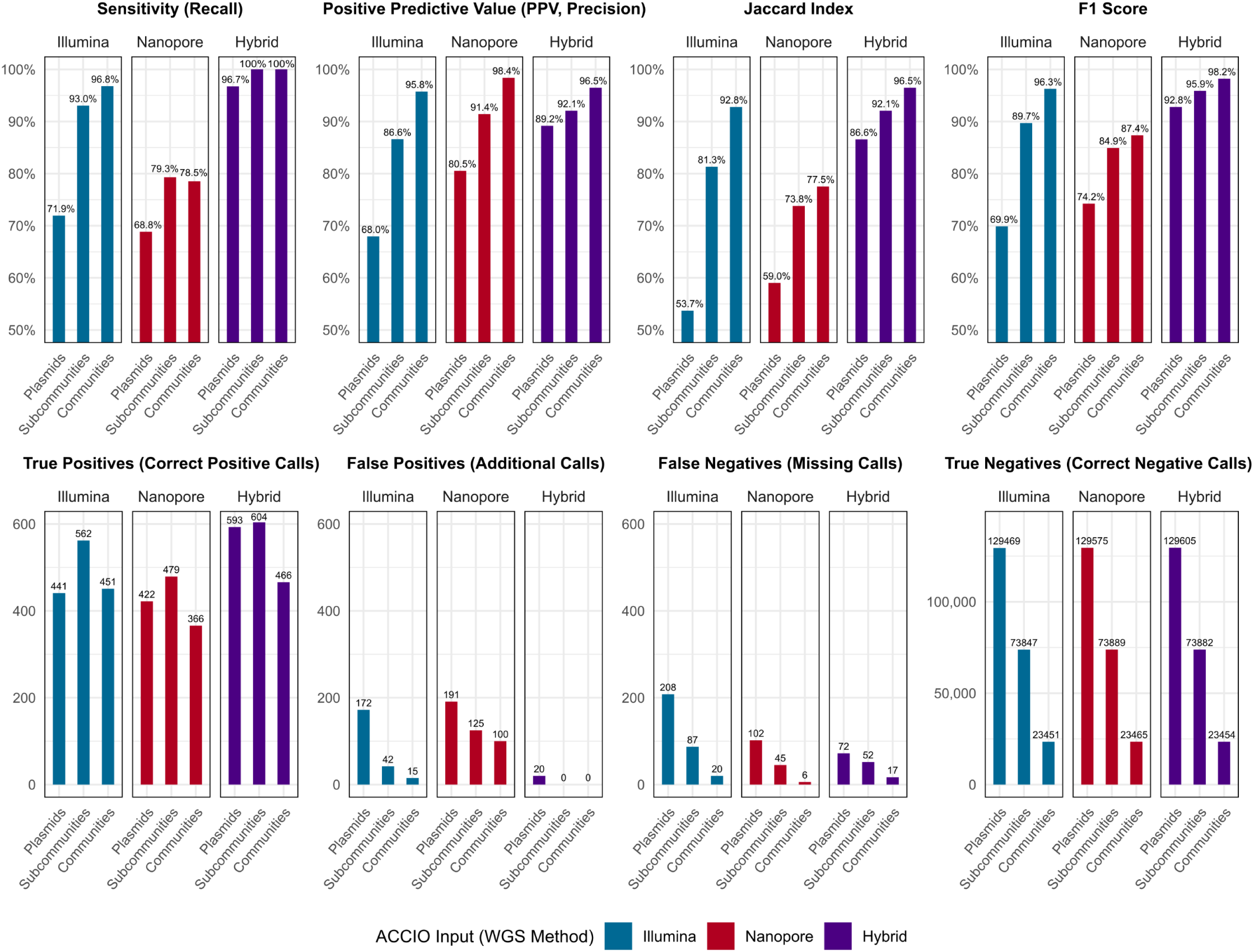
ACCIO performance by assembly input type. Classification counts (True Positives, False Positives, False Negatives, True Negatives) and performance metrics (Sensitivity, Positive Predictive Value (PPV), Jaccard Index, F1 score) for ACCIO calls generated using three whole genome sequencing assembly type inputs (Illumina, Nanopore, & Hybrid) were calculated with respect to the “ground-truth” set of plasmids (and their corresponding plasmid subcommunities and plasmid communities) recovered from study isolates as circular-and-complete plasmid contigs ≥10kb in length with identifiable plasmid replicon hits (≥80% similarity; [% identity * % coverage] / 100) in the PlasmidFinder database.

**Table 1.**
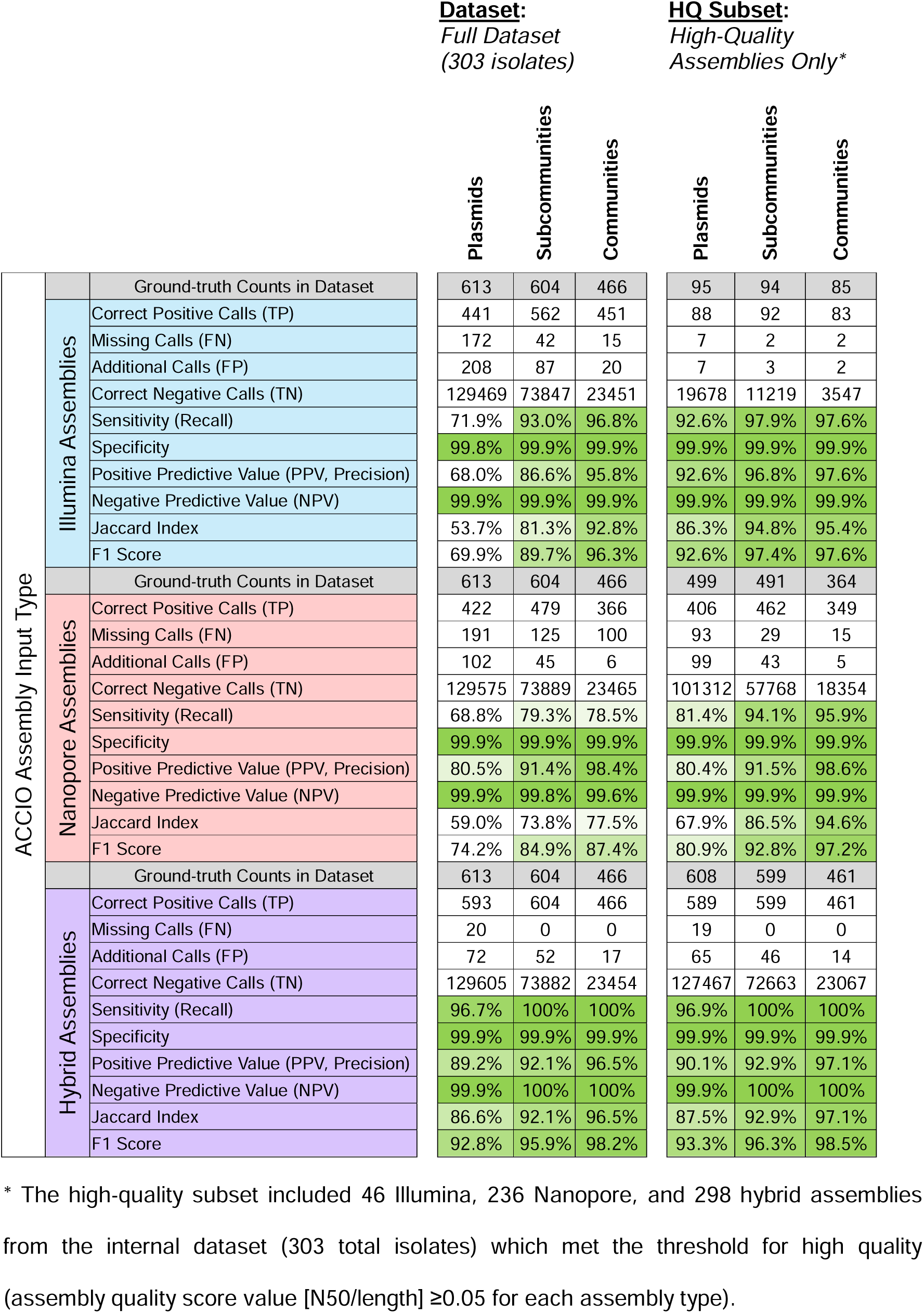
ACCIO performance by assembly input type and assembly quality.

#### Performance using hybrid assembly inputs

ACCIO achieved the highest performance metrics when using hybrid assemblies as inputs. At the plasmid level, ACCIO performed with a sensitivity of 96.7% and PPV of 89.2% (F1 = 92.8%), and at the subcommunity and community levels, it had no missing calls (FN = 0, 100% sensitivity), minimal additional calls (FP = 52 subcommunities, FP = 17 communities), and PPVs of 92.1% and 96.5% (F1 scores 95.9% and 98.2%) at the subcommunity and community levels, respectively. Of the three assembly types, hybrid assemblies are the most ‘complete’, typically resolving plasmid structures as entire, circular contigs (32,35). Thus, in hybrid assemblies, ACCIO’s plasmid-level missed calls are attributed to situations where highly similar plasmids in the database were chosen over an exact match—an issue that resolved when evaluating at the subcommunity level.

#### Plasmid, subcommunity, and community level differences

When plasmid content is assessed at the subcommunity level, ACCIO collapses highly related plasmids into single groupings using Pling, which is meant to capture plasmids that are similar enough to suggest recent common ancestry (41). Expressing calls at the subcommunity level instead of the plasmid level provides enough flexibility to resolve the ambiguity among highly related database plasmids while still maintaining the resolution required to flag groups of plasmids that could plausibly represent recent clonal transmission or horizontal transfer events. While the highest overall performance for every assembly type is seen when evaluating plasmid content at the community level (i.e. F1 scores of 98.2%, 96.3%, and 87.4% for hybrid, Illumina, and Nanopore assemblies, respectively), even when using the relatively conservative threshold of <0.3 containment distance, plasmid communities still represent a much broader landscape of relatedness, consisting of plasmids that are related enough to contain large proportions of shared sequence but not necessarily indicating recent common ancestry that would be relevant to epidemiology. A limitation of the present study is a lack of sufficient evidence to confirm whether subcommunities truly represent putative clusters of horizontal plasmid transfer. Future work will prioritize investigating plausible epidemiological linkages between plasmids in subcommunities created at different thresholds of genetic relatedness to provide more definitive evidence of an appropriate clustering threshold.

#### Performance using Illumina- and Nanopore-only assembly inputs

Performance when using Illumina and Nanopore assemblies largely followed a similar pattern to hybrid results; sensitivity and PPV markedly improved when moving from the plasmid level to subcommunity and community levels; when using Illumina-only input, ACCIO achieved a plasmid-level sensitivity of 71.9% and PPV of 68.0% (F1 = 69.9%), which rose to 93.0% sensitivity and 86.6% PPV (F1 = 89.7%) at the subcommunity level. Nanopore input showed similar performance, with a plasmid-level sensitivity of 68.8% and PPV of 80.5% (F1 = 74.2%), which rose to a sensitivity and PPV of 79.3% and 91.4% (F1 = 84.9%), respectively, at the subcommunity level. Despite similar overall performance at the subcommunity-level between Illumina and Nanopore inputs (F1 differed by < 5 percentage points; 89.7% vs 84.9%), Illumina resulted in fewer missed calls at the cost of more additional calls (FN = 42, FP = 87), while Nanopore input resulted in comparatively more missing calls and fewer additional calls (FN = 125, FP = 45). This pattern was consistently observed between Illumina and Nanopore results across all plasmid and plasmid grouping levels.

#### Average errors per isolate and error rates in context

Across virtually all assembly types and grouping levels, ACCIO maintained specificities and NPV’s > 99.9%. In practice, ACCIO assesses for the presence or absence of each reference database plasmid in each input isolate assembly, however, each validation set isolate harbored only a handful of plasmids, while the database contained hundreds to check against. Consequently, this evaluation design floods the contingency table with true negatives, and the number of additional and missing calls can become very low relative to the total number of true negative calls made per isolate, an imbalance which can cause specificity and NPV to appear inflated. Therefore, to better gauge the practical impact of these errors, we also report FP and FN both as an average per isolate and as a rate per 10,000 TN calls (**Table S1**). At the plasmid level, Illumina input averaged 0.69 FP and 0.57 FN calls per isolate, or 16.1 FP and 13.3 FN calls for every 10,000 TN calls. Importantly, these errors were not spread uniformly across isolates; over half of isolates were completely free of false positives (53.7%, 163/303) or false negatives (56.4%, 171/303). Nanopore showed a comparable rate of missed calls, but roughly half as many extra calls (FP = 0.34), while hybrid assemblies had the lowest error rates. When evaluating at the subcommunity level, error rates for Illumina and Nanopore assemblies dropped substantially, and errors became concentrated to fewer problematic isolates; for example, Illumina fell to just 0.29 FP and 0.14 FN per isolate on average, meaning false positives fell by ∼60% and false negatives by ∼75% compared with the plasmid level, and 77.9% (236/303) and 88.8% (269/303) of isolates were now without FP and FN, respectively. Thus, we regard ACCIO as generally conservative, especially at the subcommunity level, and its additional and missed calls are moderately rare, considering that hundreds of database plasmids are checked against each genome. Jaccard Index provides a measure of overall performance between the two assembly types which is particularly resistant to inflation by the large number of true negatives present in the dataset; Illumina assemblies had a substantially higher Jaccard Index at the subcommunity level (81.3% vs 73.8%), which suggests that, despite more additional calls, Illumina assemblies better recovered the same set of truly present plasmid subcommunities seen in hybrid assemblies.

#### Explaining short-versus long-read differences

The trends observed between Illumina and Nanopore performance could be explained, in part, by instances where the shorter contigs of fragmented short-read assemblies produce more partial matches across database plasmids, inflating the number of additional calls while ultimately having fewer missed calls. Long-read-only assemblers sometimes fail to assemble plasmids, particularly when they are small, and can be prone to misassembly (erroneous placement of plasmids into chromosomes or chimeric mis-joins) (31). Because our reference database excludes small plasmids, the lower sensitivity seen in Nanopore data is more likely attributed to assembly artifacts, low coverage, or poor assembly quality. While the longer contigs of long-read-only assemblies help to reduce the spurious matches to database plasmids that produce false positives, they also appear to be more susceptible to mis-joins or inaccuracies that reduce the quality of contig matches to ground-truth plasmids. The latter, which use short reads for draft construction and long reads for bridging, may produce plasmids whose final structures align more closely with multiple fragmented Illumina contigs than with a single Nanopore contig. A limitation of the present study is that we only formally evaluated three assembly pipelines which produced the best results in our experience: SPAdes / Unicycler (Illumina), Flye (Nanopore), and Unicycler (hybrid). We also evaluated plasmidSPAdes for Illumina data, a specialized assembler for plasmids, though performance was lower than that of the base SPAdes / Unicycler assemblies, likely due to over-filtering when designating non-plasmidic contigs and instances where sequences from distinct replicons were merged (data not shown) (57,58). As ACCIO detects plasmids using assembled contigs, both sequencing type and assembler choice ultimately influence ACCIO’s accuracy. Benchmarking ACCIO’s performance using additional long- and short-read assemblers is a priority for future work.

#### Performance by assembly quality

In addition to assembler, assembly quality is a key determinant of ACCIO’s performance. Analysis using a subset of high-quality validation dataset isolate genomes, each passing a quality threshold of [N50/length] ≥ 0.05, showed large gains in performance, particularly for Illumina and Nanopore data (**Table 1**. HQ Subset). At the plasmid level, Illumina performance improved markedly in the high-quality subset; from an F1 score of 69.9% (71.9% sensitivity, 68.0% PPV) in the full dataset to 92.6% (92.6% sensitivity, 92.6% PPV) in the high-quality set. At the subcommunity level, Illumina data showed more modest gains (change in F1 from 93.0% to 97.9%), but this improvement brought performance to near-perfect levels; when restricted to high-quality assemblies, Illumina input achieved performance on par with that of hybrid assemblies (which only displayed slight improvement in the high-quality set), showing minimal differences (∼1 percentage point) in F1 score across all groupings. Using high-quality Nanopore assemblies also resulted in marked increases to sensitivity and F1 despite very stable PPVs; we observed 51.3% (FN = 191 to 93) and 76.8% (FN = 125 to 29) reductions in missing calls at the plasmid and subcommunity levels, respectively. The sizeable change in false negatives between sets suggests that low-quality Nanopore assemblies contributed disproportionately to missed calls. It is also important to note that only a small fraction of Illumina assemblies (15.2%, 46/303), a majority of Nanopore assemblies (77.9%, 236/303), and nearly all hybrid assemblies (98.3%, 298/303) met the quality threshold for inclusion into this sub-analysis. We therefore recommend that best practice with ACCIO involves being aware of the quality of the input genomes, as assemblies meeting or exceeding the quality threshold [N50/length] ≥ 0.05 showed improved performance across the board. Enforcing this cutoff appears especially practical for Nanopore and hybrid assemblies, as we were able to retain the majority of isolates while improving performance. Considering our QC thresholds (35× for Illumina and 10× for Nanopore), systematically requiring higher coverage (e.g., ≥50× for short-read and ≥100× for long-read assemblies) and/or optimizing assembly parameters could reasonably increase the proportion of high-quality assemblies (44,50,59). While ACCIO can be applied to assemblies of any quality, users should interpret the results for isolates below the quality threshold with more caution, as the likelihood of both false negative and false positive calls is increased.

### 7.3 Performance assessment on three external datasets

#### Generalizability and variable datasets

To assess ACCIO’s generalizability in datasets other than our own internal validation set, we evaluated its performance using three external datasets which varied in size, plasmid content, and species composition (**Table 2**). All three datasets included isolates with hybrid assemblies available which were used to generate a clustered plasmid database, determine a validation set of isolates with ≥1 plasmid, and define the ground-truth set of plasmids for each validation set isolate. When available, we retrieved assemblies directly from NCBI or ENA and otherwise obtained raw reads to construct Illumina and/or Nanopore assemblies using the same methods described above.

**Table 2.**
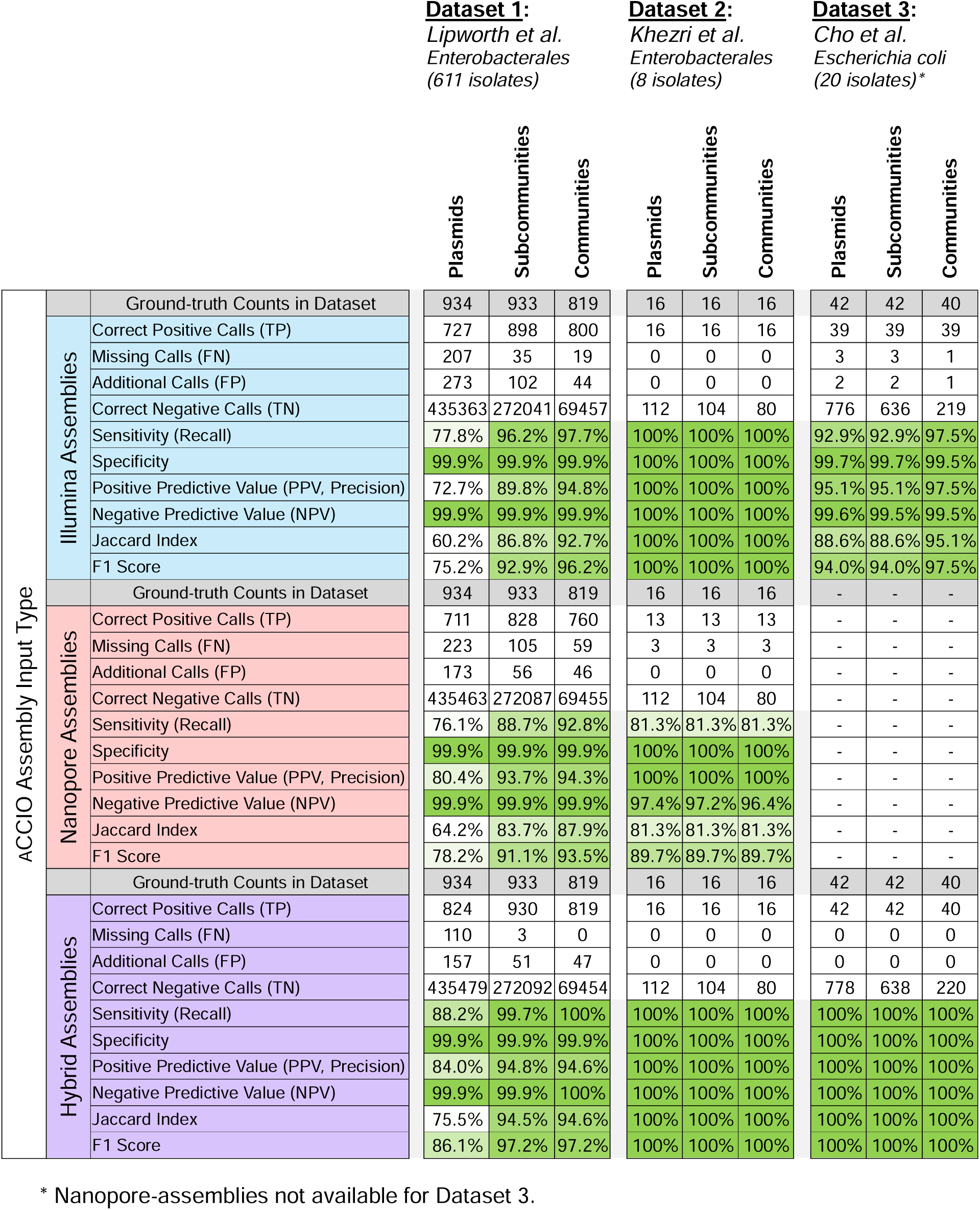
ACCIO performance in external datasets by assembly input type.

#### External dataset 1

**Lipworth et al** (60). This large-scale dataset included a total of 738 Enterobacterales hybrid assemblies which were screened for plasmids. Initially, 1880 circular candidate plasmid sequences were identified by ACCIO, 1135 (60.4%) of which were removed as duplicates due to size filtering and/or lack of a plasmid replicon. The total number of representative plasmid sequences in the final database created by ACCIO was 745 plasmids from 611 validation set isolates. The size of this validation dataset, along with its similar species composition and larger plasmid database, provided an opportunity to evaluate ACCIO on a dataset similar to our own but collected in a distinct geographic setting. Performance using this external set was comparable to the results observed in our internal analysis; in fact, at the subcommunity level, the overall performance using Illumina and Nanopore input (F1 = 92.9% and F1 = 91.1%, respectively) was higher than the performance observed using our own internal validation set. In this dataset, the performances of Illumina and of Nanopore were comparable to one another, reaching sensitivities of 96.2% and 88.7% and PPVs of 89.8% and 93.7% for Illumina and Nanopore, respectively. The Jaccard Index for Illumina and Nanopore were also comparable at each grouping, typically remaining within a difference of 3 percentage points or less. Their error profiles were largely the same as observed in our internal set; Nanopore tended to produce fewer additional calls at the cost of more missing calls, with the opposite being true for Illumina. Taken together, these results provide evidence supporting ACCIO’s generalizability and suitability for application in other research or surveillance settings.

#### External dataset 2

**Khezri et al** (35). This small set of Enterobacterales contained 9 hybrid assemblies which were screened for plasmids. A total of 44 candidate plasmid sequences were identified by ACCIO, 28 (63.6%) of which were removed during filtering and deduplication, leaving a total of 16 representative plasmids from 8 validation set isolates in the final database, less than 5% of the size of our internal database (340 plasmids). This set was chosen to emulate the use of ACCIO during a small-scale investigation, where the database might only be populated with a few plasmids of interest. Across all grouping levels, ACCIO performed with perfect precision and recall when using Illumina or hybrid assemblies, correctly identifying all 16 plasmids amongst the isolates. At each grouping level, Nanopore input missed 3 calls (81.3% sensitivity and 97% PPV) but did not make any additional calls, resulting in an F1 score of 89.7% for all groupings. These results demonstrate that ACCIO performs well when looking for a few pertinent plasmids from a small set of isolates, especially in Illumina and hybrid data.

#### External dataset 3

**Cho et al** (61). This set of *E. coli* ST131 (Nanopore assemblies unavailable) contained 21 isolates with hybrid assemblies from which 130 candidate plasmids were identified. After 89 plasmids were removed during filtering and deduplication, a total of 41 plasmids from 20 validation set isolates remained in the final database. As this set is comprised of isolates from a single lineage of *E. coli*, it was selected as a test of ACCIO’s ability to differentiate and accurately call plasmids within more closely related isolates. Performance was strong across all groupings, with Illumina F1 scores at 94.0% or higher. For Illumina, at both the plasmid and subcommunity levels, there were a total of 3 false negative and 2 false positive calls. Hybrid assemblies achieved perfect agreement with the set of ground-truth plasmids. These results suggest that ACCIO can distinguish plasmids even among closely related isolates using hybrid assemblies and, in a case like this, Illumina input is a viable alternative with comparable performance.

### 7.4 Additional benchmarking and considerations

#### Comparison of ACCIO with a current standard toolkit (38)

To contextualize ACCIO’s results, we compared them against those obtained using a common plasmid reconstruction and clustering toolkit, MOB-suite, applied to the same internal validation dataset. MOB-suite represents a widely adopted and highly useful standard for plasmid typing and clustering; using a string of its modules, MOB-cluster, MOB-recon, and MOB-typer, and the addition of Pling clustering, we obtained plasmid, subcommunity, and community calls which could be directly compared to calls from ACCIO. Across nearly all input types and grouping levels, ACCIO significantly outperformed MOB-suite in both sensitivity and precision, with higher F1 scores for all combinations of sequencing type and plasmid groupings (**Fig. 5**, **Table 3**). For example, at the plasmid and subcommunity level, ACCIO demonstrated significantly higher sensitivity and PPV for Illumina (*p*□<L0.001 for both), Nanopore (*p*L<L0.01 and *p*L<L0.05, respectively), and hybrid input (*p*L<L0.001 for both). The only exception was at the community level with Illumina assemblies, where MOB-suite exhibited slightly higher sensitivity (98.8% vs 96.8%, *p* < 0.05), suggesting that, when using Illumina input assemblies with the goal of identifying the most correct community calls, MOB-suite offers a slightly higher sensitivity at the cost of more additional calls (45 vs 135) and subsequently lower F1 score (92.1% vs 96.3%). The lower performance of MOB-suite can likely be attributed to two factors: 1) it chooses a nearest neighbor database plasmid exclusively based on mash distance, whereas ACCIO considers multiple other additional measures of relatedness, and 2) it relies on error-prone plasmid reconstruction as part of MOB-recon. A recent benchmarking analysis attempted to reconstruct plasmids from the Illumina assemblies of plasmid-harboring *E. coli* using six different plasmid reconstruction tools, and found MOB-suite able to reconstruct only half of the plasmids found by hybrid assembly, despite that being the best performance out of all the tools evaluated (58). In addition, it was noted that, at best, less than 30% of plasmids carrying ARGs could be correctly reconstructed by MOB-suite and large, ARG-harboring plasmids reconstructed by MOB-suite contained chromosomal contamination 40% of the time. These results highlight circumventing plasmid reconstruction when calling database plasmids as a main advantage of ACCIO.

**Fig. 5.**
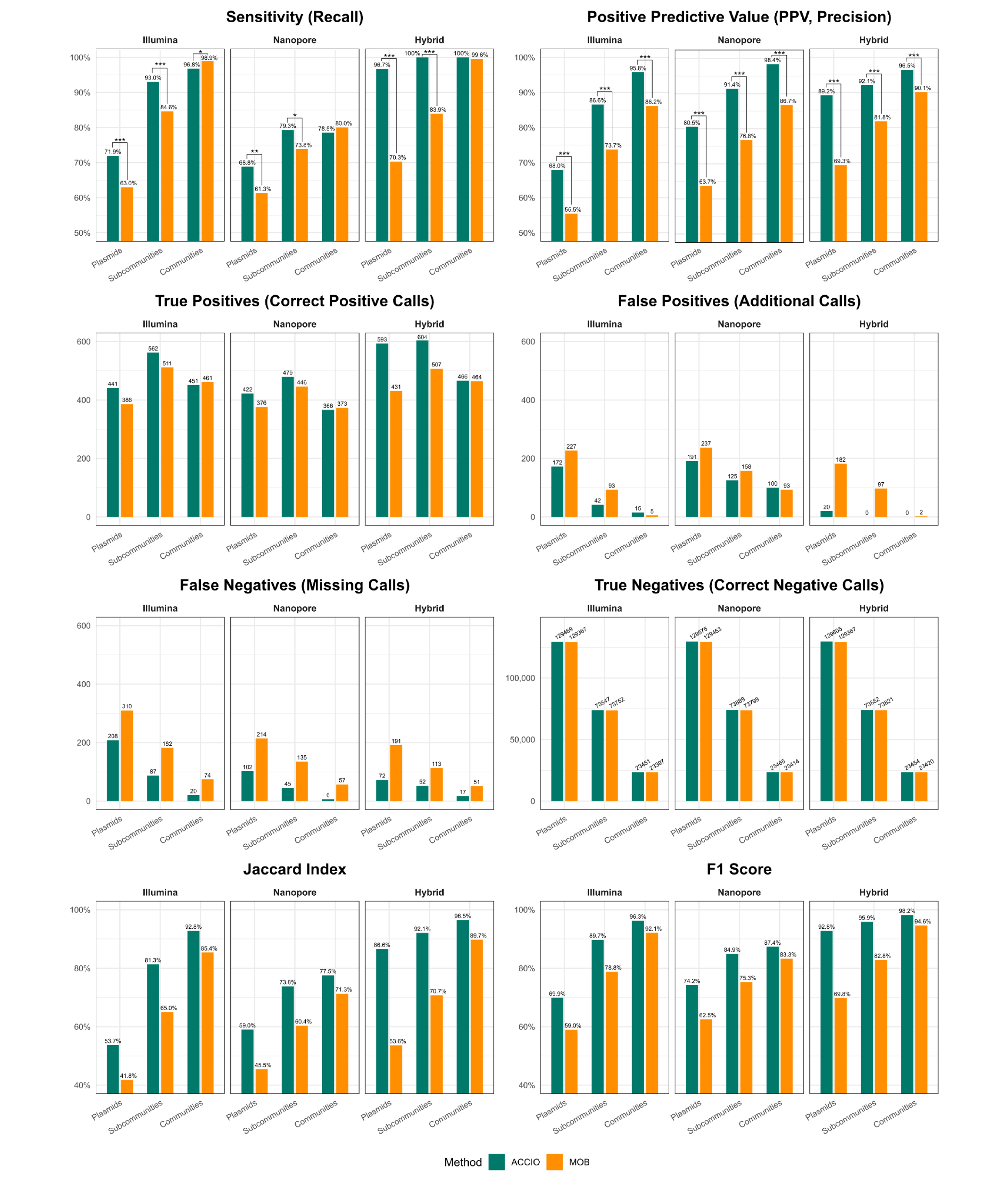
Performance of ACCIO and MOB-suite across assembly input types relative to ground-truth set. Classification counts (True Positives, False Positives, False Negatives, True Negatives) and performance metrics (Sensitivity, Positive Predictive Value (PPV), Jaccard Index, F1 score) for both ACCIO and MOB-suite calls generated using three whole genome sequencing assembly type inputs (Illumina, Nanopore, & Hybrid) were calculated with respect to the “ground-truth” set of plasmids (and their corresponding plasmid subcommunities and plasmid communities) recovered from study isolates as circular-and-complete plasmid contigs ≥10kb in length with identifiable plasmid replicon hits (≥80% similarity; [% identity * % coverage] / 100) in the PlasmidFinder database. Significance values for two-sample tests for equality of proportions using chi-square approximations are shown over connecters for relevant panels (no asterisk, p<0.1; *, p<0.05; **, p<0.01; ***, p<0.001).

**Table 3.**
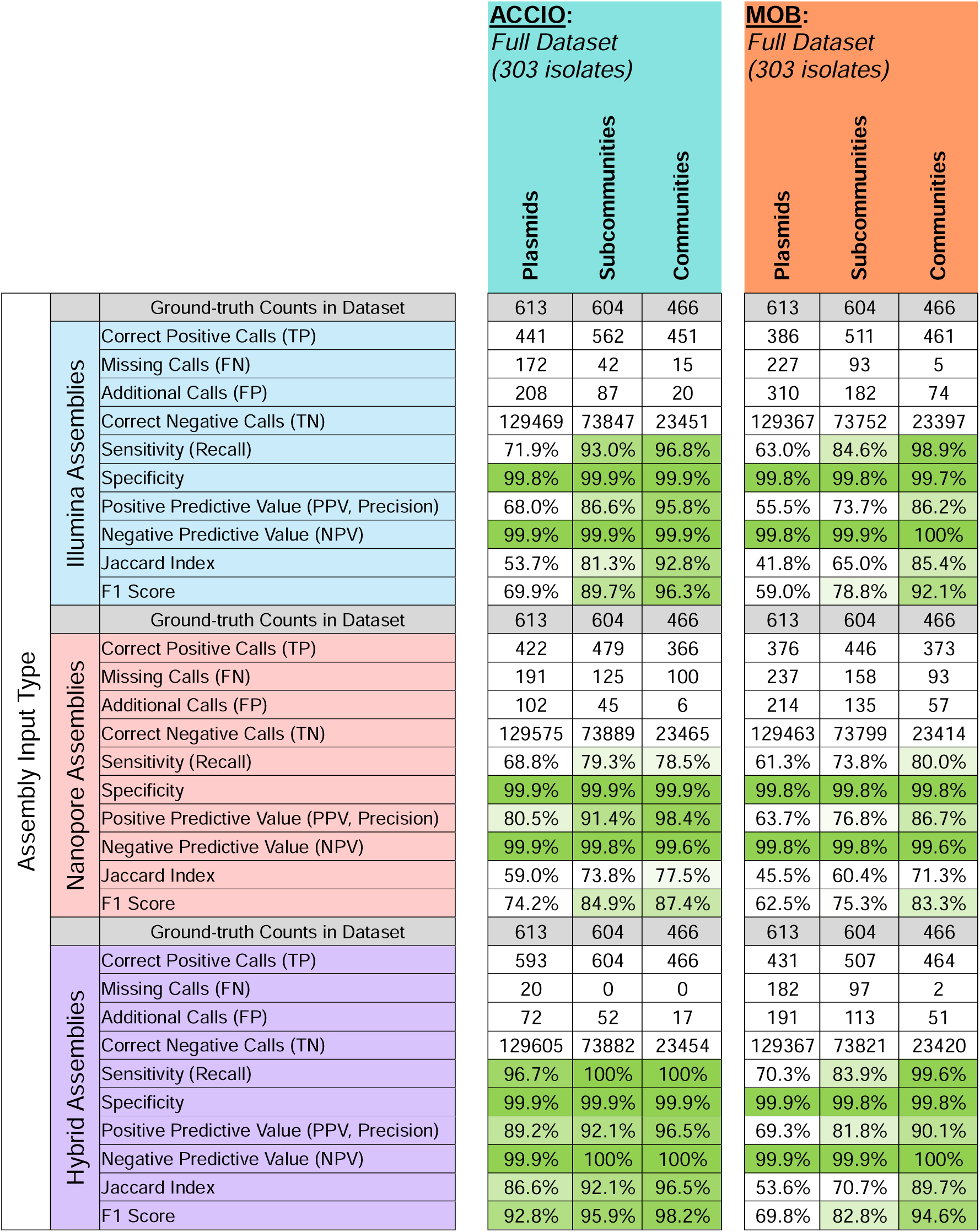
Comparison of ACCIO vs MOB performance across assembly input types.

#### Novel plasmid discovery

Assembly contigs suspected to be of plasmid origin remaining after plasmid calling are used in reconstruction to identify the presence of putative novel plasmids not yet represented in the local database. Across all three assembly types for the 303-isolate validation set, ACCIO identified 152 total instances of novel plasmids (length range 10,039–1,152,119 bp): 11 in Illumina assemblies, 96 in Nanopore assemblies, and 45 in hybrid assemblies. However, not all plasmidic contigs identified were utilized during reconstruction—122, 148, and 50 isolates had at least one unused plasmidic contig for Illumina, Nanopore, and hybrid assemblies, respectively. While these findings suggest that longer and more complete assembly contigs appear to improve novel plasmid recovery, any novel plasmids identified should be regarded solely as putative and used primarily to help target sequencing when a plasmid is expected but not present in the database plasmid calls.

#### Ground-truth agreement with base-assembly replicons

Plasmids which were truly present in a genome—but not circularized in hybrid assemblies—would not be identified as part of the ground-truth set or included in the plasmid database creation step by ACCIO. This represents an inherent limitation when defining the ground-truth set of plasmids for each isolate. In the instance an isolate truly contains a plasmid which was not circularized in the hybrid assembly, and a genetically similar plasmid is represented in the database (either from another isolate or supplemented sequence), the present study design penalizes this as a false positive, when it instead represents a limitation of the gold-standard method for recovery of plasmids. We conducted a supplemental analysis to measure whether the set of plasmid replicons identified in the base assemblies of the internal validation isolates had stronger overlap with the set of plasmids called by ACCIO or the full set of ground-truth plasmids recovered from hybrid assemblies (**Fig. S4**, **Table S2**). We identified a total of 1279, 1116, and 1280 plasmid replicons in the base Illumina, Nanopore, and hybrid assemblies, respectively. Across all assembly types, the set of replicons in the plasmids called by ACCIO had greater overlap with those found in the base assemblies than the ground-truth set. Jaccard Index values were higher for ACCIO than for the ground-truth set in Illumina (71.5% vs 69.0%), Nanopore (67.6% vs 58.9%), and hybrid data (75.1% vs 70.0%), indicating ACCIO had a closer agreement with base-assembly replicons, with the differences observed for Nanopore and hybrid comparisons being more apparent. The large number of additional calls observed in the ground-truth set for Nanopore assemblies may be explained, in part, by assembler-driven differences between long-read-only and hybrid assembly contigs. When looking at hybrid assemblies, there were 66 more missing calls in the ground-truth set compared to the ACCIO set, suggesting that ACCIO calls some plasmids whose replicons are present in the base assembly, but may not have been circularized in the hybrid assembly. Accordingly, throughout this study, we also refer to false positives as additional calls, recognizing that this limitation of the gold standard precludes a more definitive classification.

#### Secondary validation using ACCIO hybrid plasmids as a ground-truth

Using the hybrid assemblies of all 500 study isolates screened to create the plasmid database, we defined the set of plasmids called as present by ACCIO as the new ground-truth set of plasmids. Trends in these results mirrored the primary analysis; Illumina captured more correct positive calls at the expense of more extra calls and Nanopore was more conservative at the expense of more missed calls (**Fig. S5**, **Table S3**). Because the ground-truth in this analysis is derived from ACCIO itself, these results should be interpreted as an internal comparison rather than an estimate for external accuracy. Overall, it appeared that the Illumina calls had more overlap with hybrid than hybrid did with Nanopore.

#### Reads vs no reads

We conducted a supplementary analysis to determine the optimal default settings regarding ACCIO’s use of reads (with or without reads) (**Fig. S6**, **Table S4**). Results showed marginal improvements (≤*0.2 percentage points*) when using Nanopore and hybrid input without reads. However, the analysis also showed substantial increases in performance (sensitivity 93.0% vs 85.3%) when using reads vs no reads, respectively, for Illumina data. This boost in performance is likely due to calculation of a gap score only for Illumina isolates, as the gap score was intended to use reads to help measure the coverage of regions of the database plasmids not covered by fragmented matching contigs. As such, the default settings of ACCIO use reads for Illumina input and no reads for Nanopore and hybrid input.

#### Computational time and memory

We benchmarked computational time and memory using a random subset of 54 bacterial isolates from nine species, including *K. pneumoniae* (n=29), *E. coli* (n=6), *E. hormaechei* (n=6), *E. faecium* (VRE, n=5), methicillin-resistant *Staphylococcus aureus* (MRSA, n=2), *Clostridiodes difficile* (n=2), *Klebsiella oxytoca* (n=2), *Klebsiella variicola* (n=1), and *P. mirabilis* (n=1). Analyses were performed on an in-house server (Dell PowerEdge T640; 172 TB storage, 1.48 TB RAM, dual Intel Xeon Gold 6240R CPUs). ACCIO’s mean runtime per sample was 10.6 min (range 0.9–34.7) with a mean peak memory use of 4.6 GB RSS (± 0.7 GB). Species with higher plasmid content, such as *Klebsiella* spp., required longer runtimes than those with few or no plasmids, such as *C. difficile*.

### 7.5 Conclusions

We developed and evaluated ACCIO’s ability to infer plasmid content using a large internal validation dataset consisting of clinical and surveillance hospital isolates and three external datasets of varying size and composition. We then benchmarked its performance against the widely-used MOB-suite toolkit. We found that ACCIO successfully leverages the information stored in assembly contigs, regardless of assembly type, to infer which plasmids and plasmid groupings are present in an isolate. While performance was consistently strong, results also showed that missed calls and additional calls can occur to varying degrees, particularly when using Illumina-only or Nanopore-only assembly input at the plasmid level. In order of relative importance, interpretation of ACCIO results should consider three key factors: 1) assembly type (and assembler), 2) grouping level, and 3) assembly quality. We have shown that each of these can influence detection accuracy and error rates, and therefore the level of confidence in the resulting plasmid, subcommunity, or community calls made by ACCIO. Based on our results in external datasets, ACCIO is a versatile tool which could be generalized to a variety of sequencing platforms, datasets, and settings. Although ACCIO was meant to help circumvent the need for additional sequencing and hybrid assembly whenever possible, as part of a real-world epidemiologic surveillance pipeline, ACCIO’s use is ideally supplemented with periodic hybrid assemblies or other analyses functioning as a spot checker for instances where a plasmid is suspected but not called. More broadly, ACCIO provides a tool to quickly and accurately detect important plasmids and plasmid groupings from routine WGS data, offering researchers and public health or hospital surveillance programs a practical means to identify and monitor potential plasmid-mediated resistance without the need for additional sequencing. In future work, we aim to apply ACCIO to other datasets to investigate plasmid epidemiology at varying geographic scales. We also plan to integrate ACCIO into our existing real-time genomic surveillance system to characterize plasmid epidemiology across thousands of hospital isolates sequenced with Illumina technology, with the integration strategy requiring careful consideration.

## Supporting information

Supplementary_Data_S1_Isolate_and_Plasmid_Metadata

Supplementary_Data_S2_ACCIO_Performance_Sheets

## Data Availability

Short- and long-read sequencing data have been deposited in the NCBI Sequence Read Archive (SRA) under multiple BioProjects, and corresponding hybrid genome assemblies are available in GenBank. Accession numbers for all BioProjects, BioSamples, and SRA datasets are provided in Supplementary Data S1. All supporting data, software code, and experimental/analysis protocols are provided within the article, in supplementary data files, or on GitHub.

https://github.com/mpgriffith/accio

## Repositories

All source code, supplementary data, and sequencing data associated with this study are openly available: the ACCIO source code can be accessed on GitHub at https://github.com/mpgriffith/accio. Short- and long-read sequencing data have been deposited in the NCBI Sequence Read Archive (SRA) under multiple BioProjects, and corresponding hybrid genome assemblies are available in GenBank. Accession numbers for all BioProjects, BioSamples, and SRA datasets are provided in Supplementary Data S1.

## Supplementary Data

Supplementary Data S1. Isolate and Plasmid Metadata

Supplementary Data S2. ACCIO Performance Sheets

## 9. Author statements

### 9.1 Author contributions

***NJR:*** *Conceptualization; Data curation; Formal analysis; Investigation; Methodology; Project Administration; Software; Validation; Visualization; Writing – original draft; Writing – review and editing*.

***MPG:*** *Data curation; Formal analysis; Investigation; Methodology; Software; Validation; Writing – review & editing*.

***VRS:*** *Conceptualization; Visualization; Writing – review & editing*.

***KDW:*** *Resources; Writing – review & editing*.

***AJS:*** *Funding acquisition; Writing – review & editing*.

***LLP:*** *Resources; Funding acquisition; Writing – review & editing*.

***GMS:*** *Supervision; Funding acquisition; Writing – review & editing*.

***MMB:*** *Supervision; Formal analysis; Writing – review & editing*.

***DVT:*** *Supervision; Conceptualization; Resources; Methodology; Funding acquisition; Writing – original draft; Writing – review & editing*.

***LHH:*** *Supervision; Conceptualization; Resources; Project Administration; Funding acquisition; Writing – original draft; Writing – review & editing*.

### 9.4 Ethical approval

Ethics approval was obtained from the University of Pittsburgh Institutional Review Board (Protocol STUDY21040126).

## 9.5 Acknowledgements

Special thanks to Yohei Doi, MD, PhD, and Ryan Shields, PharmD, MS, for providing select samples included in the study, Jane Marsh, PhD, for her role in developing the EDS-HAT infrastructure, and Vaughn Cooper, PhD, for whole genome sequencing assistance. The name ACCIO (ack-ee-oh) was chosen for its Latin meaning, “I summon,” reflecting a tool designed to call plasmids out of assemblies, and a spell many readers might recognize, too.

## 8. Figures and Tables

**Fig S1.**
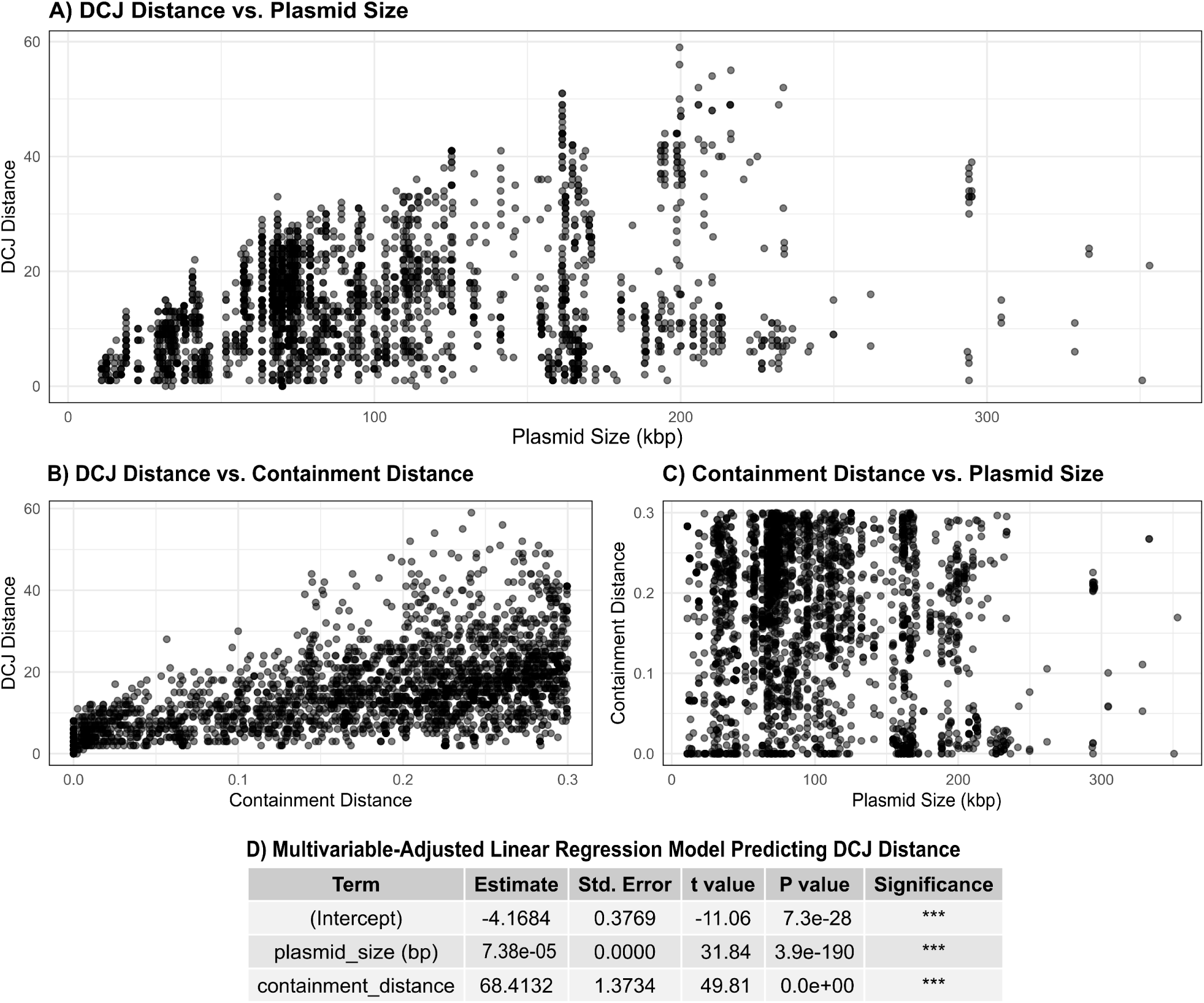
Relationships between plasmid size, containment distance, and DCJ-Indel (DCJ) distance among pairwise plasmid comparisons. **A)** DCJ distance is plotted against the size of the smaller plasmid in each pairwise comparison of plasmids in our local database with containment distance <0.3; **B)** Containment distance shows a positive correlation with DCJ distance. **C)** No apparent correlation was observed between smaller plasmid size and containment distance, suggesting that containment distance likely explains some variation in DCJ distance independently of plasmid size. **D)** A linear regression model was fit to predict DCJ with size of the smaller plasmid, adjusting for containment distance. Holding containment distance constant, we observed that 1) when the smaller plasmid size is ∼120 kb, the predicted DCJ distance is ∼4, and 2) conservatively, for every ∼20 kb increase in the size of the smaller plasmid beyond 120kb, the DCJ distance increases by ∼1.

**Fig S2.**
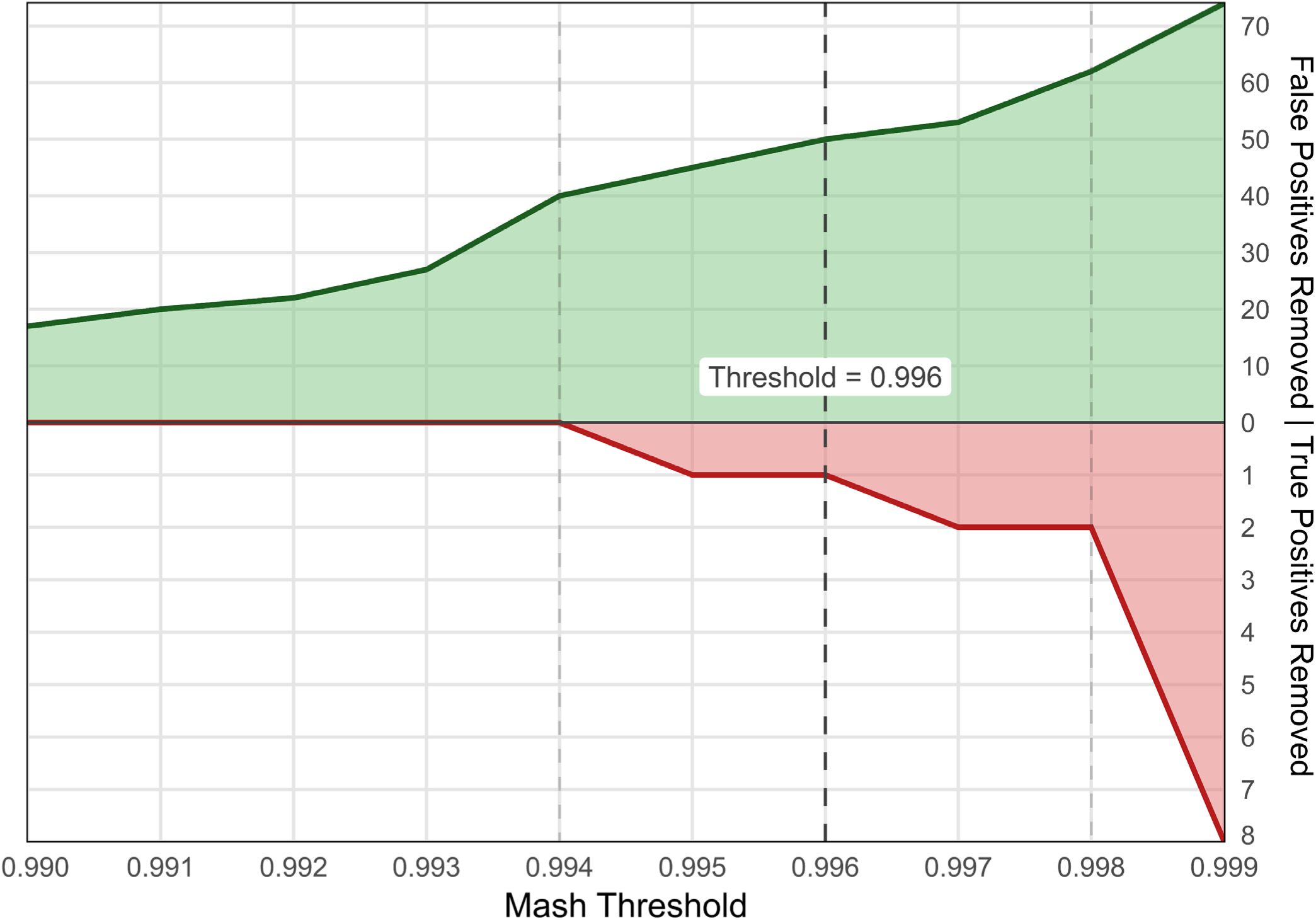
Optimization for mash score threshold. The number of false positives (green) and true positives (red) removed at each mash score thresholds is shown. ACCIO default threshold: 0.996 (dark dashed line); alternative threshold range: 0.994-0.998 (light dashed lines).

**Fig. S3.**
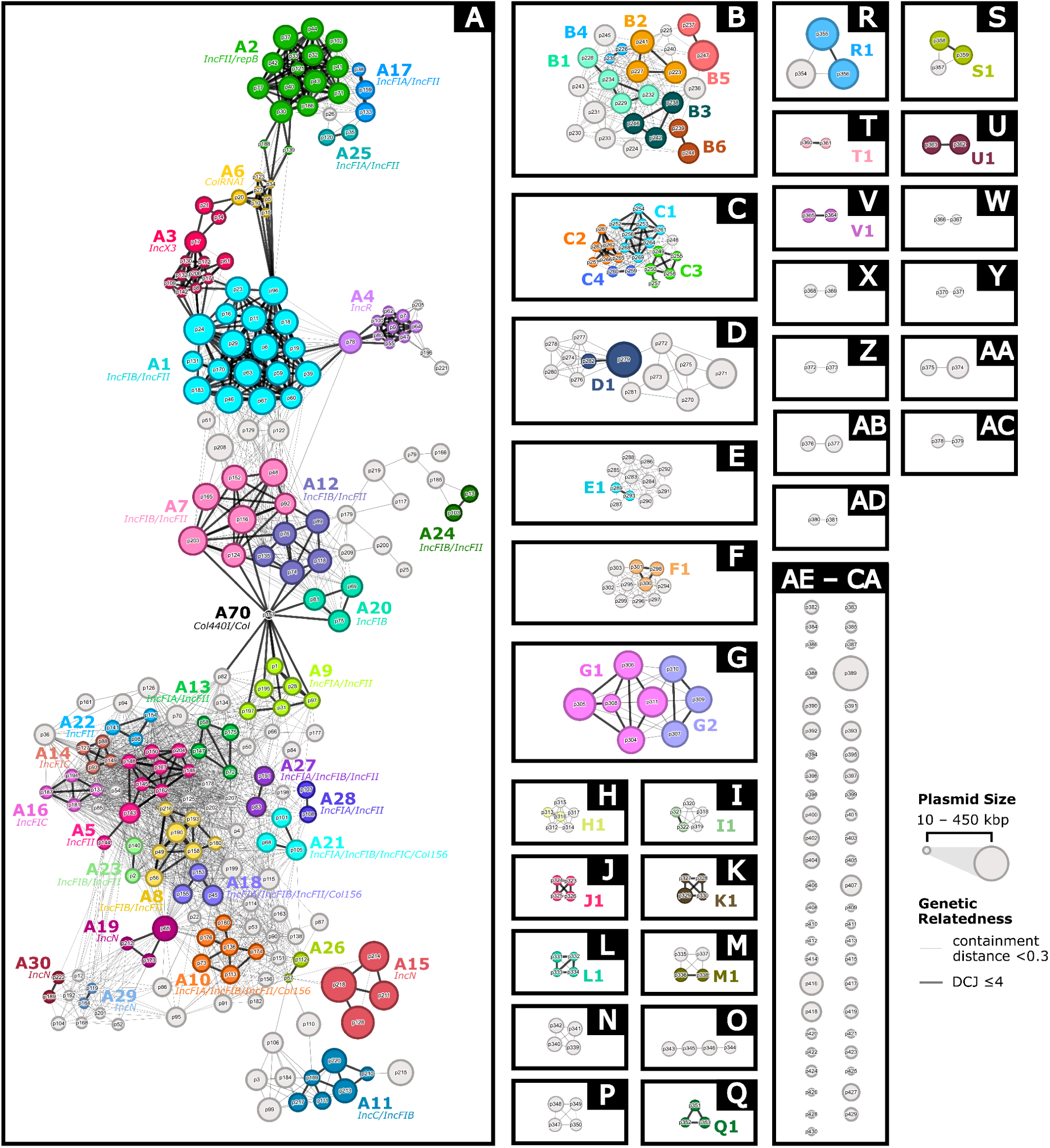
Clustered local reference plasmid database created using ACCIO (plasmid numbers labeled). The local reference plasmid database (n=430 representative plasmids) was clustered using Pling to define plasmid communities (boxed, labeled A–CA by descending size) based on a containment distance threshold of <0.3. Plasmid study numbers are labeled p1–p430 in chronological order of host isolate culture date within each community. Within each community, subcommunities (colored groups, labeled as community letter plus subcommunity number for all subcommunities containing ≥2 plasmids) represent groups of closely related plasmids defined by high structural similarity (DCJ-Indel distance ≤4 for plasmids <120 kbp, +1 DCJ per additional 20 kbp). Light grey edges represent links between plasmids below the containment distance threshold; black edges represent links between plasmids whose containment distance and DCJ distance are both below the thresholds for relatedness. Plasmid nodes are scaled by size (∼10–450 kbp). Light grey nodes represent plasmids which lacked strong relatedness to any other plasmid in the database and thus were assigned to their own subcommunity. A black node denotes a hub plasmid found in Community A, treated here as its own subcommunity. The plasmid replicons encoded by the majority of plasmids in each subcommunity are listed for Community A.

**Fig. S4.**
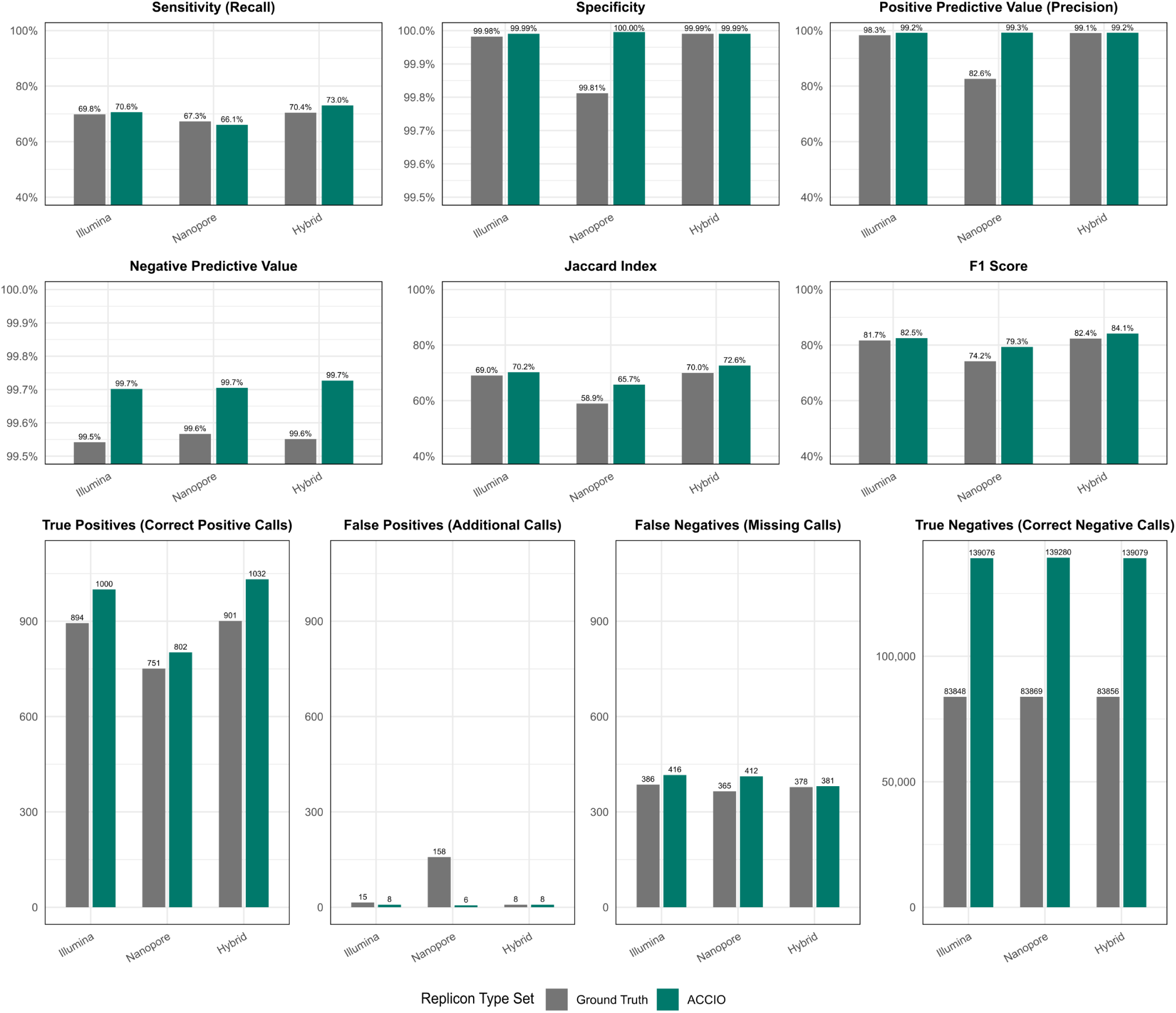
Performance of ACCIO and ground-truth replicon calls against base assembly replicons. Classification counts (True Positives, False Positives, False Negatives, True Negatives) and performance metrics (Sensitivity, Positive Predictive Value [PPV], Jaccard Index, F1 score) were calculated for the set of plasmid replicons in the set of plasmids called by ACCIO and the set of plasmid replicons in the ground-truth plasmid set against the set of plasmid replicons present in the base assembly (identifiable plasmid replicon hits in the base isolate assembly with ≥80% similarity; [% identity * % coverage] / 100 in the PlasmidFinder database) as the reference. Results are shown across three base assembly types (Illumina, Nanopore, Hybrid).

**Fig. S5.**
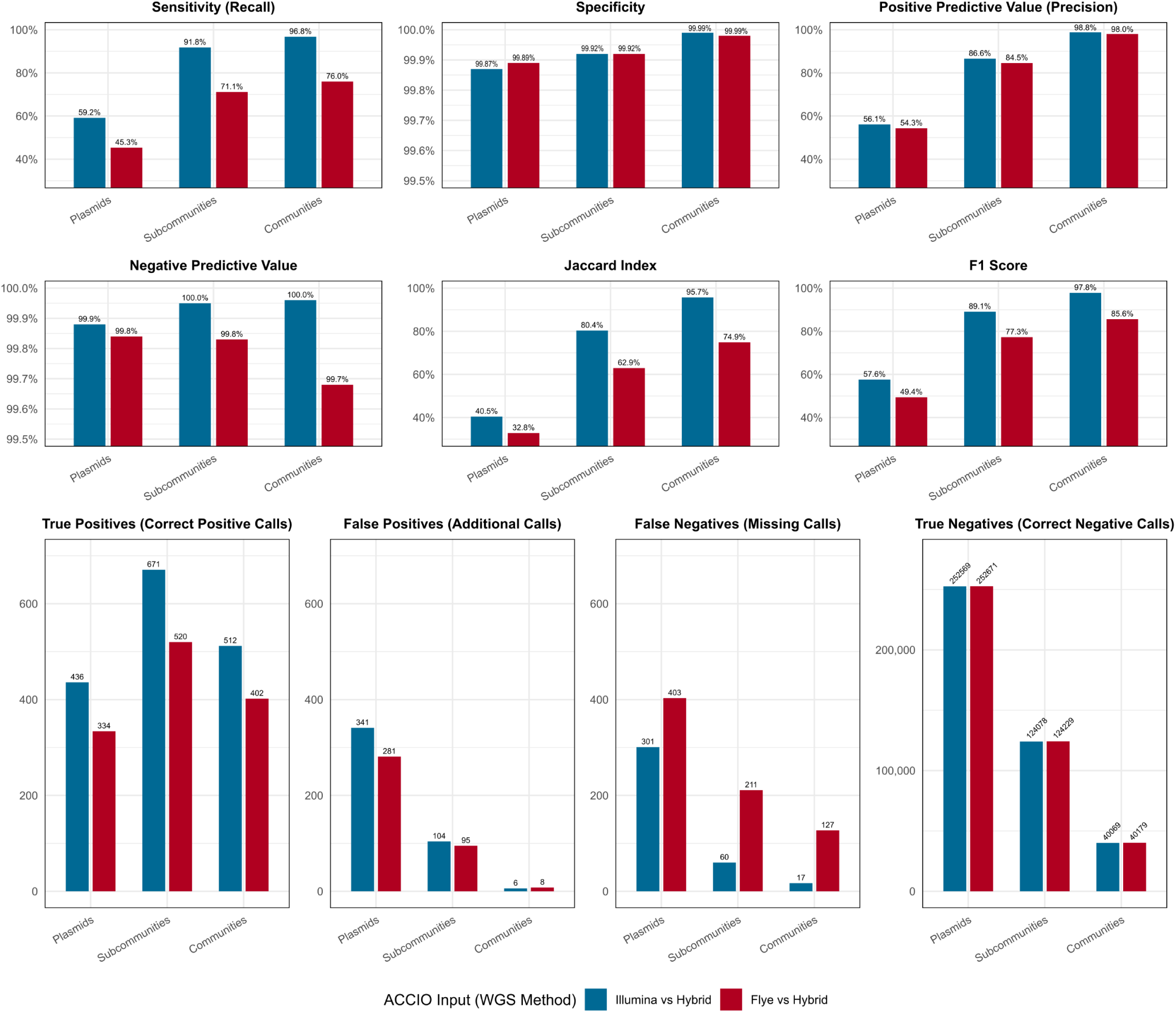
Performance of ACCIO with Illumina and Nanopore inputs compared to ACCIO Hybrid. Classification counts (True Positives, False Positives, False Negatives, True Negatives) and performance metrics (Sensitivity, Positive Predictive Value (PPV), Jaccard Index, F1 score) for ACCIO calls generated using two whole genome sequencing assembly type inputs (Illumina & Nanopore) were calculated with respect to the set of plasmids, plasmid subcommunities, and plasmid communities called as present by ACCIO using hybrid assembly inputs.

**Fig. S6.**
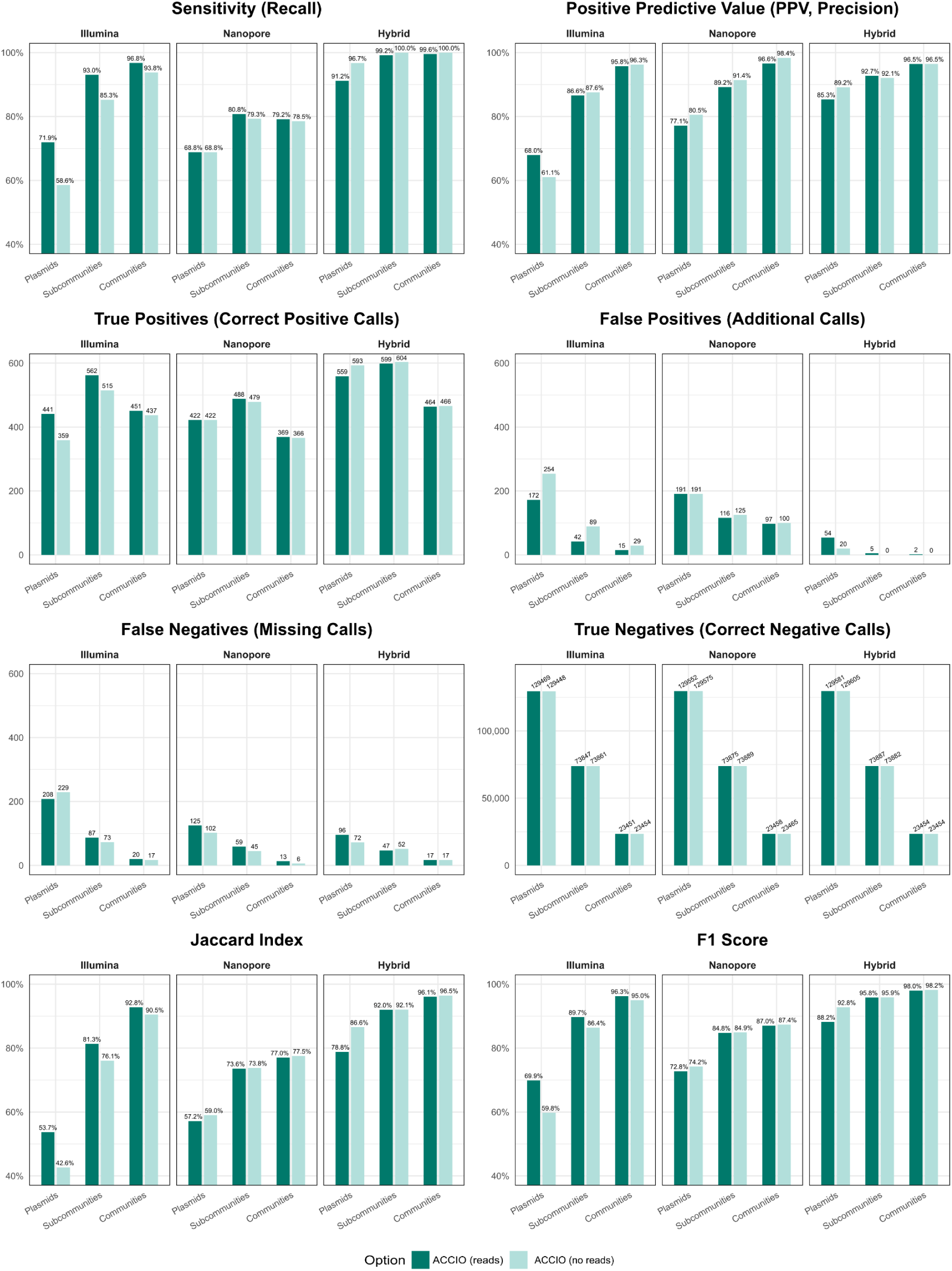
Performance of ACCIO with and without using reads across assembly input types relative to ground-truth set. Classification counts (True Positives, False Positives, False Negatives, True Negatives) and performance metrics (Sensitivity, Positive Predictive Value (PPV), Jaccard Index, F1 score) for ACCIO calls generated using three whole genome sequencing assembly type inputs (Illumina, Nanopore, & Hybrid) with and without the use of sequencing reads were calculated with respect to the “ground-truth” set of plasmids (and their corresponding plasmid subcommunities and plasmid communities) recovered from study isolates as circular-and-complete plasmid contigs ≥10kb in length with identifiable plasmid replicon hits (≥80% similarity; [% identity * % coverage] / 100) in the PlasmidFinder database.

**Table S1.**
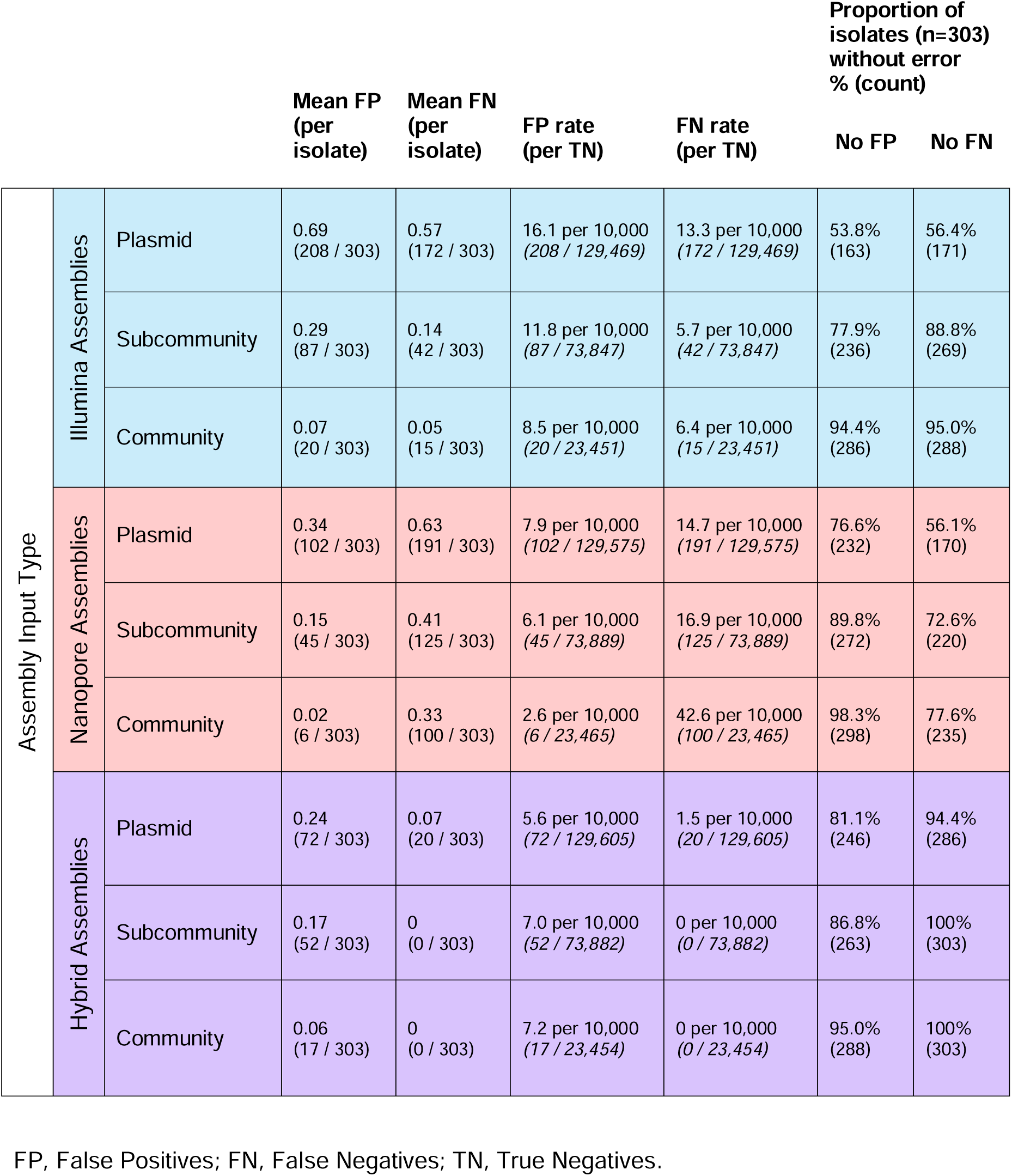
ACCIO error rates by assembly type.

**Table S2.**
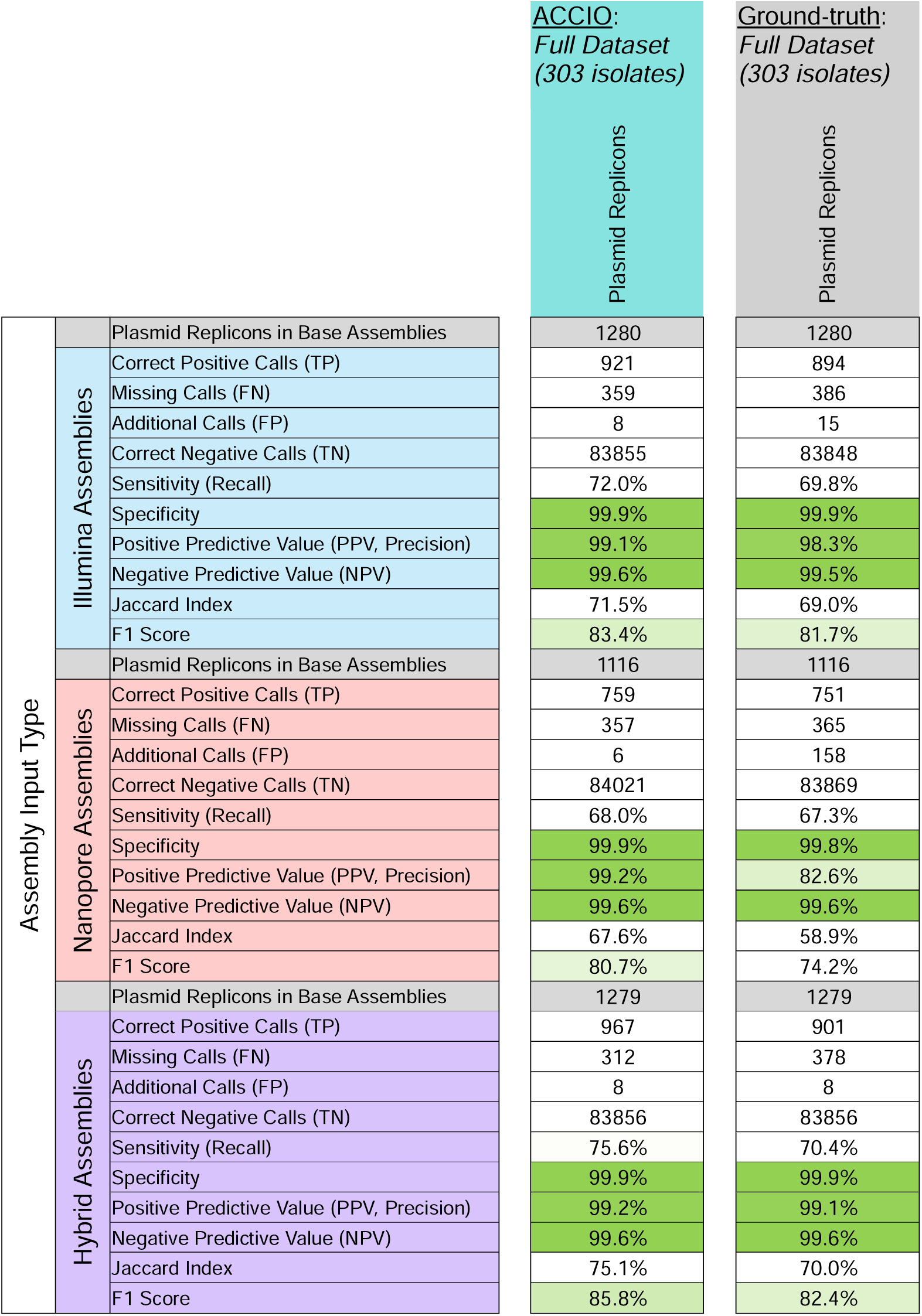
Agreement with replicons present in base assemblies: replicons in the ‘Ground-truth’ plasmid set versus those in plasmids called by ACCIO.

**Table S3.** Internal validation using ACCIO hybrid plasmid calls as ground-truth.

Dataset: Full Dataset (500 isolates)
|  |  |  | Plasmids | Subcommunities | Communities |
| --- | --- | --- | --- | --- | --- |
| ACCIO Assembly Input Type | Illumina Assemblies | Ground-truth Counts in Dataset | 705 | 694 | 514 |
|  |  | Correct Positive Calls (TP) | 479 | 630 | 495 |
|  |  | Missing Calls (FN) | 226 | 64 | 19 |
|  |  | Additional Calls (FP) | 218 | 65 | 11 |
|  |  | Correct Negative Calls (TN) | 214077 | 122241 | 38975 |
|  |  | Sensitivity (Recall) | 67.9% | 90.8% | 96.3% |
|  |  | Specificity | 99.9% | 99.9% | 99.9% |
|  |  | Positive Predictive Value (PPV, Precision) | 68.7% | 90.6% | 97.8% |
|  |  | Negative Predictive Value (NPV) | 99.9% | 99.9% | 99.9% |
|  |  | Jaccard Index | 51.9% | 83.0% | 94.3% |
|  |  | F1 Score | 68.3% | 90.7% | 97.1% |
|  | Nanopore Assemblies | Ground-truth Counts in Dataset | 705 | 694 | 514 |
|  |  | Correct Positive Calls (TP) | 429 | 510 | 382 |
|  |  | Missing Calls (FN) | 276 | 184 | 132 |
|  |  | Additional Calls (FP) | 120 | 39 | 6 |
|  |  | Correct Negative Calls (TN) | 214175 | 122267 | 38980 |
|  |  | Sensitivity (Recall) | 60.9% | 73.5% | 74.3% |
|  |  | Specificity | 99.9% | 99.9% | 99.9% |
|  |  | Positive Predictive Value (PPV, Precision) | 78.1% | 92.9% | 98.5% |
|  |  | Negative Predictive Value (NPV) | 99.9% | 99.8% | 99.7% |
|  |  | Jaccard Index | 52.0% | 69.6% | 73.5% |
|  |  | F1 Score | 68.4% | 82.1% | 84.7% |

**Table S4.**
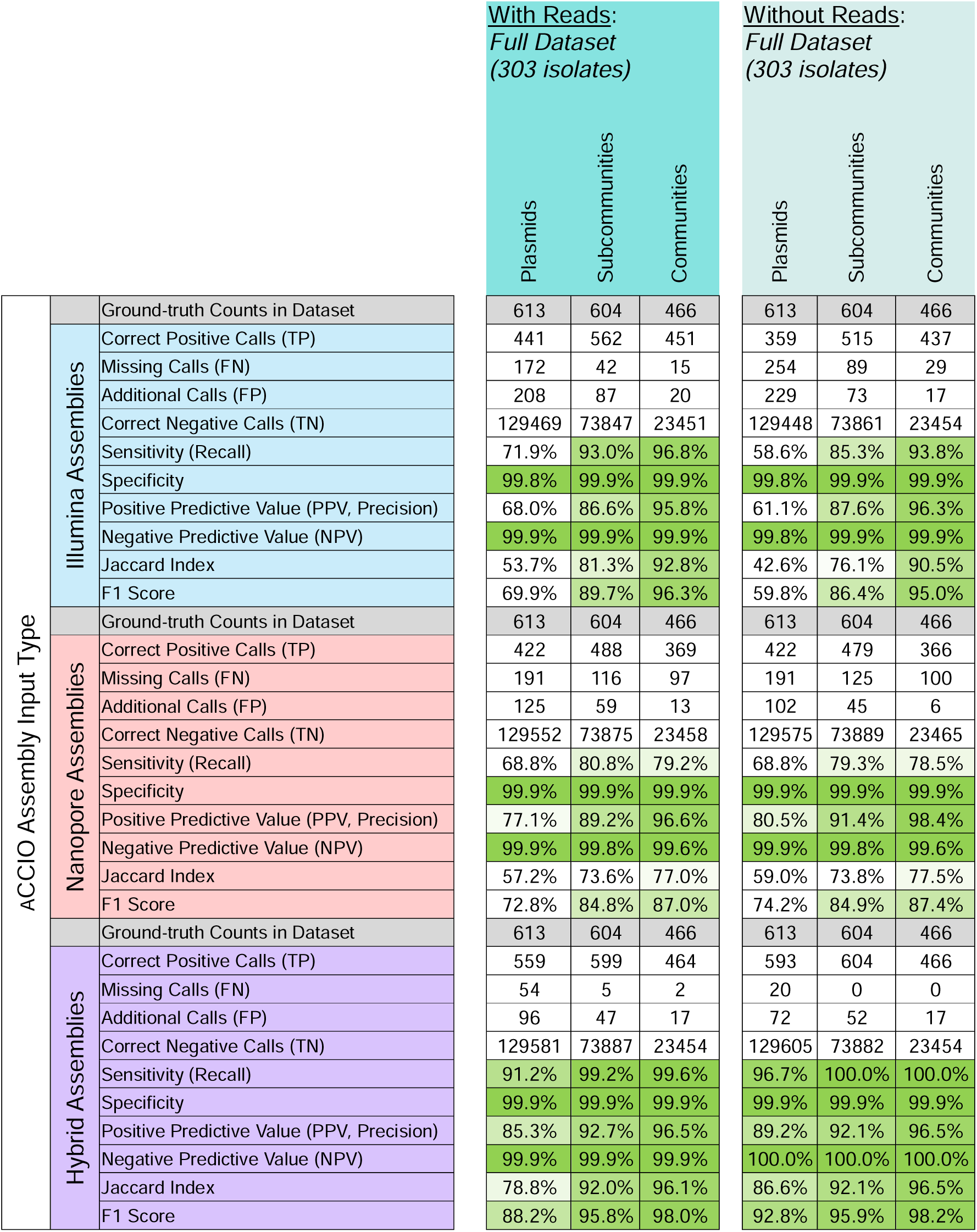
ACCIO performance with and without sequencing reads.

